# Risk Factors for SARS-CoV-2 Seropositivity in a Health Care Worker Population

**DOI:** 10.1101/2020.12.17.20248430

**Authors:** Sebastian D. Schubl, Cesar Figueroa, Anton M. Palma, Rafael R. de Assis, Aarti Jain, Rie Nakajima, Alguimantas Jasinkas, Danielle Brabender, Ariana Naaseh, Oscar Hernandez Dominguez, Ava Runge, Shannon Skochko, Justine Chinn, Adam James Kelsey, Kieu Thai Lai, Weian Zhao, Peter Horvath, Delia Tifrea, Areg Grigorian, Abran Gonzales, Suzanne Adelsohn, Frank Zaldivar, Robert Edwards, Alpesh N. Amin, Michael J. Stamos, Philip S. Barie, Philip L. Felgner, Saahir Khan

**Affiliations:** Department of Surgery, University of California Irvine Health, Orange, CA; Institute for Clinical and Translational Sciences, University of California Irvine, Irvine, CA; Department of Physiology and Biophysics, University of California Irvine, Irvine, CA; School of Medicine, University of California Irvine, Irvine, CA; Department of Pharmaceutical Sciences, University of California Irvine, Irvine, CA; Department of Pathology, University of California Irvine Health, Orange, CA; Department of Surgery, University of Southern California, Los Angeles, CA; Department of Medicine, University of California Irvine Health, Orange, CA; Departments of Surgery and Medicine, Weill Cornell Medicine, New York, NY; Department of Medicine, University of Southern California, Los Angeles, CA

**Keywords:** Coronavirus disease 2019, Severe acute respiratory syndrome coronavirus-2, health care worker, antibody, hospital epidemiology

## Abstract

**Background:** Protecting health care workers (HCWs) during the coronavirus disease 2019 (COVID-19) pandemic is essential. Serologic testing can identify HCWs who had minimally symptomatic severe acute respiratory syndrome coronavirus-2 (SARS-CoV-2) infections that were missed by occupational screening based on daily symptom and temperature checks. Recent studies report conflicting results regarding the impact of occupational factors on SARS-CoV-2 seropositivity amongst HCWs.

**Methods:** The study population included all hospital workers at an academic medical center in Orange County, California. SARS-CoV-2 seropositivity was assessed from a fingerstick blood specimen using a coronavirus antigen microarray, which compares IgM and IgG antibodies against a panel of SARS-CoV-2 antigens with positive and negative controls to identify prior SARS-CoV-2 infection with 98% specificity and 93% sensitivity. Demographic, occupational, and clinical factors were surveyed and their effect on seropositivity estimated using multivariable logistic regression analysis.

**Results:** Amongst 1,557 HCWs with complete data, SARS-CoV-2 seropositivity was 10.8%. Risk factors for increased seropositivity included male gender, exposure to COVID-19 outside of work, working in food or environmental services, and working in COVID-19 units. Amongst the 1,103 HCW who were seropositive but missed by occupational screening, additional risk factors included younger age and working in administration.

**Conclusions:** SARS-CoV-2 seropositivity is significantly higher than reported case counts even amongst HCWs who are meticulously screened. Seropositive HCWs missed by occupational screening were more likely to be younger, work roles without direct patient care, or have COVID-19 exposure outside of work.

**Key Points:** SARS-CoV-2 seropositivity risk factors amongst health care workers included male gender, nonoccupational exposure, food or environmental services role, and COVID-19 unit location. Those missed by occupational screening were younger, in roles without direct patient care, or exposed outside of work.

## Background

Protecting health care workers (HCWs) during the coronavirus disease 2019 (COVID-19) pandemic is essential to mounting an effective response, as outbreaks among this population could potentially cripple health care delivery. Current case identification relies on symptom and temperature screening with follow-up testing by severe acute respiratory syndrome coronavirus-2 (SARS-CoV-2) reverse transcriptase-polymerase chain reaction (rt-PCR). This approach underestimates disease prevalence due to the high proportion of asymptomatic infections and the impaired sensitivity of rt-PCR due to suboptimal timing, flawed specimen collection, over-amplification, or low viral load[1, 2]. Given the importance of asymptomatic persons in the dissemination of SARS-CoV-2, identification of risk factors that may augment identification of asymptomatic infection in HCWs is crucial to protecting patients and the health care system[1].

Serologic testing can help to determine the true cumulative prevalence of COVID-19 by identifying previously infected persons who had minimal symptoms so were missed by the current testing paradigm[3, 4]. Multiple COVID-19 seroprevalence studies have been performed in different populations but are limited by low specificity in low-prevalence populations or potential selection bias from the use of convenience sampling[5-10]. Estimated seroprevalence among HCW varies widely, with some studies finding similar or even lower prevalence compared to the surrounding community[7, 11-14].

Risk factors for SARS-CoV-2 infection amongst HCW were initially extrapolated from studies of hospitalized patients with severe disease that may not be generalizable to the population at large[15]. Early studies to identify risk factors amongst HCWs were performed in early outbreak setting prior to current infection control practices so may not be currently applicable[16, 17]. More recent studies have conflicting results as to whether occupational exposures confer an increased risk of SARS-CoV-2 seropositivity and are limited by heterogeneity and suboptimal performance of the assays and inadequate control for confounding[18, 19].

This study measured SARS-CoV-2 seropositivity amongst 1,557 HCWs at the University of California-Irvine Health, a 418-bed academic medical center in Orange County, California, from May 15th to June 30th 2020, using a novel coronavirus antigen microarray (CoVAM). This CoVAM utilizes 11 SARS-CoV-2 antigens to determine prior infection with 98% specificity and 93% sensitivity based on validation in 91 rt-PCR-positive cases and 88 pre-pandemic negative controls[20]. This performance and level of validation compares favorably to other serologic assays based on a single antigen[21-23]. A digital survey collected data on potential demographic, occupational, and clinical risk factors for SARS-CoV-2 infection. A multivariable analysis tested the null hypothesis that demographic, clinical, and occupational risk factors do not affect SARS-CoV-2 seropositivity among HCWs.

## Methods

### Study Design, Setting, and Population

The study was approved by the Institutional Review Board of the University of California-Irvine under Protocol HS 2020-5818. All employees who worked in the hospital were eligible. Universal daily symptom and temperature screening was initiated on April 14, 2020, with subsequent immediate rt-PCR testing for any HCW with symptoms or fever or disclosure of a confirmed or suspected COVID-19 contact. A primary study site in the main hospital building was open from May 15 to May 29, 2020 to all employees who provided electronic consent (open enrollment cohort). In addition, all employees who had been tested for SARS-CoV-2 by rt-PCR due to symptoms or possible exposure, or who provided direct patient care in COVID-19 clinical units or similar control units, were invited via email and provided electronic consent to participate at a secondary study open from May 15 to June 30, 2020 (targeted enrollment cohort). This second cohort was included to enrich the study for HCW with COVID-19 infection, symptoms, or exposure.

### Study Procedures

Participants were given a unique study identifier and a mobile phone link to a Research Electronic Data Capture (REDCap, Vanderbilt University, Nashville, TN) survey to collect data on demographic, clinical, and occupational risk factors (Appendix A). At the primary study site, participants then underwent capillary blood collection via fingerstick using a disposable lancet into microfuge capillary tubes. After centrifugation at 1500 x g for 10 min, supernatant plasma was collected, frozen, and transported for laboratory analysis. At the secondary study site, participants underwent phlebotomy into gold-top tubes (BD Biosciences, San Jose, CA) for centrifugation and collection of serum, from which an aliquot was frozen and transported for laboratory analysis. All specimens were labeled with unique identifiers accessible only via a secure key.

### Laboratory Assays

The CoVAM includes 67 antigens from respiratory viruses, including 11 antigens from SARS-CoV-2 (Sino Biological U.S. Inc., Wayne, PA). Antigens were printed onto microarrays in quadruplicate, probed with serum specimens and secondary antibodies for IgM and IgG, and imaged to determine background-subtracted median fluorescence intensity[24-26]. Briefly, CoVAM data for each specimen were compared with 91 rt-PCR-positive cases with blood collected ≥ 7 d (range, 7-50 d, median 11 d) post-symptom onset and 88 pre-pandemic controls with blood collected prior to November 1, 2019, which were split randomly into 70% derivation and 30% validation cohorts. Based on IgM and IgG antibodies against the 11 SARS-CoV-2 antigens on the array, a linear regression model was trained on positive and negative controls in the derivation cohort to determine optimal weighted combinations of reactive antigens to calculate composite SARS-CoV-2 antibody titers that discriminate the two groups, with reactivity thresholds selected to achieve maximum sensitivity while maintaining ≥ 98% specificity. The model was tested on the validation cohort and achieved 92.7% sensitivity and 97.7% specificity for detecting prior SARS-CoV-2 infection based on composite IgM or IgG positivity.

### Statistical Analysis

SARS-CoV-2 seropositivity was calculated as the proportion of HCWs who were classified as seropositive in the study population and within categories of each demographic, clinical and occupational risk factor, expressed as the odds ratio (OR) with 95% confidence interval compared to the study population. In order to assess the associations between clinical and occupational risk factors and seropositivity, multivariable models were constructed to control for potential confounding due to demographic and health-related factors associated with both occupational exposure and underlying risk for seropositivity. We forced demographic variables into the model *a priori*, which included age, gender, and race/ethnicity, which have well-established associations with occupation and health outcomes. Health-related covariates included comorbidities (asthma or COPD, diabetes mellitus, hypertension, self-reported smoking or vaping) and known COVID-19 exposure outside of work, which would likely influence seropositivity. Occupation-related variables included self-reported role, location, and COVID-19 patient contact. Health- and occupation-related exposures were selected based on bivariate associations with seropositivity using a p<0.1 criterion for inclusion. The final model for clinical and occupational risk factors was adjusted for age (quartiles), gender, race/ethnicity (Asian, White, Latino, Black, and Mixed/other/not reported), known COVID-19 exposure outside of work, and workplace role, location, and COVID-19 patient contact. Adjusted analyses were conducted among the entire sample and replicated among the subgroup of HCWs not tested previously via rt-PCR. Model fit was evaluated using the Hosmer-Lemeshow goodness-of-fit test and C-statistic. All analyses were conducted using R software v4.0.3 (R Consortium for Statistical Computing, Vienna, Austria).

## Results

### Study Population

From an eligible population of 5,349 HCWs, 1,841 (34.4%) consented to participate, including 1,108 in the open enrollment cohort and 733 in the targeted enrollment cohort. Of the targeted cohort, 343 had been tested by rt-PCR for COVID-19, 237 worked in a COVID-19 unit and 153 worked in a matched control unit of similar acuity. A total of 1,557 HCWs completed the survey and provided blood specimens to be analyzed by CoVAM, including 1,044 in the open enrollment cohort and 513 in the targeted cohort. Compared to the population of Orange County, the study population has a higher proportion of females and Asian race/ethnicity.

SARS-CoV-2 seropositivity was 10.8% in the overall HCW cohort (Table 1). Seropositivity was 17.7% amongst the 419 HCWs who had been tested by rt-PCR previously, and 8.0% among the 1,138 HCW who had not. Seropositivity in the targeted versus open enrollment cohorts matched closely the seropositivity among HCWs tested previously versus not tested by rt-PCR respectively, indicating that the difference between the cohorts is largely driven by rt-PCR testing. Of the 343 HCW tested by rt-PCR, 38 were PCR+ and 322 PCR-, with 36 (94.4%) of the PCR+ testing seropositive whereas 30 (9.3%) of the PCR-were seropositive, higher than the seropositivity rate of those not tested by rt-PCR (Supplementary Table 4).

**Table 1.** Association between demographic and health-related characteristics and SARS-CoV-2 seropositivity (AB prevalence) of HCW study population and subgroups segregated by prior rt-PCR testing. (COPD, chronic obstructive pulmonary disease)

| Parameter | All HCWs (n=1,557) |  |  | Not tested by rt-PCR (n=1,138) |  |  | Tested by rt-PCR (n=419) |  |  |
| --- | --- | --- | --- | --- | --- | --- | --- | --- | --- |
|  | HCWs, n (%) | COVID-19 AB prevalence, n (%) | OR (95% CI)<sup>1</sup> | HCWs, n (%) | COVID-19 AB prevalence, n (%) | OR (95% CI)<sup>1</sup> | HCWs, n (%) | COVID-19 AB prevalence, n (%) | OR (95% CI)<sup>1</sup> |
| Total | 1557 (100) | 165 (10.6) |  | 1138 (100) | 91 (8.0) |  | 419 (100) | 74 (17.7) |  |
| Age quartiles (y) |  |  |  |  |  |  |  |  |  |
| 18-31 | 418 (26.8) | 49 (11.7) | 1.17 (0.82-1.66) | 311 (27.3) | 33 (10.6) | 1.57 (1.00-2.45) | 107 (25.5) | 16 (15.0) | 0.77 (0.41-1.38) |
| 32-38 | 382 (24.5) | 41 (10.7) | 1.02 (0.69-1.47) | 257 (22.6) | 22 (8.6) | 1.10 (0.65-1.79) | 125 (29.8) | 19 (15.2) | 0.78 (0.43-1.36) |
| 39-43 | 377 (24.2) | 35 (9.3) | 0.83 (0.55-1.21) | 275 (24.2) | 15 (5.5) | 0.60 (0.33-1.03) | 102 (24.3) | 20 (19.6) | 1.19 (0.66-2.07) |
| 49-73 | 380 (24.4) | 40 (10.5) | 0.99 (0.67-1.43) | 295 (25.9) | 21 (7.1) | 0.85 (0.50-1.38) | 85 (20.3) | 19 (22.4) | 1.46 (0.80-2.59) |
| Gender |  |  |  |  |  |  |  |  |  |
| Female | 1073 (68.9) | 100 (9.3) | 0.66 (0.48-0.93) | 781 (68.6) | 57 (7.3) | 0.75 (0.48-1.18) | 292 (69.7) | 43 (14.7) | 0.53 (0.32-0.90) |
| Male | 482 (31.0) | 64 (13.3) | 1.48 (1.05-2.06) | 355 (31.2) | 33 (9.3) | 1.28 (0.81-1.99) | 127 (30.3) | 31 (24.4) | 1.87 (1.11-3.13) |
| Other<sup>2</sup> | 2 (0.1) | 1 (50.0) | - | 2 (0.2) | 1 (50.0) | - | 0 (0) | - | - |
| Race/ethnicity |  |  |  |  |  |  |  |  |  |
| Asian | 608 (39.0) | 70 (11.5) | 1.17 (0.84-1.62) | 415 (36.5) | 26 (6.3) | 0.68 (0.42-1.07) | 193 (46.1) | 44 (22.8) | 1.93 (1.16-3.24) |
| White | 457 (29.4) | 46 (10.1) | 0.92 (0.64-1.31) | 336 (29.5) | 32 (9.5) | 1.33 (0.84-2.07) | 121 (28.9) | 14 (11.6) | 0.52 (0.27-0.94) |
| Latino | 286 (18.4) | 27 (9.4) | 0.86 (0.54-1.30) | 232 (20.4) | 18 (7.8) | 0.96 (0.55-1.61) | 54 (12.9) | 9 (16.7) | 0.92 (0.41-1.90) |
| Black | 29 (1.9) | 3 (10.3) | 0.97 (0.23-2.80) | 25 (2.2) | 2 (8.0) | 1.00 (0.16-3.46) | 4 (1.0) | 1 (25.0) | 1.56 (0.08-12.39) |
| Mixed/Other/Not reported | 177 (11.4) | 19 (10.7) | 1.02 (0.60-1.65) | 130 (11.4) | 13 (10.0) | 1.32 (0.68-2.38) | 47 (11.2) | 6 (12.8) | 0.65 (0.24-1.50) |
| Comorbidities |  |  |  |  |  |  |  |  |  |
| Any comorbidities | 370 (23.8) | 41 (11.1) | 1.07 (0.73-1.54) | 258 (22.7) | 18 (7.0) | 0.83 (0.47-1.39) | 112 (26.7) | 23 (20.5) | 1.30 (0.74-2.22) |
| Asthma or COPD | 155 (10.0) | 16 (10.3) | 0.97 (0.54-1.62) | 114 (10.0) | 9 (7.9) | 0.98 (0.45-1.92) | 41 (9.8) | 7 (17.1) | 0.96 (0.38-2.13) |
| Diabetes mellitus | 67 (4.3) | 10 (14.9) | 1.51 (0.71-2.89) | 45 (4.0) | 3 (6.7) | 0.82 (0.19-2.30) | 22 (5.3) | 7 (31.8) | 2.30 (0.85-5.68) |
| Hypertension | 172 (11.0) | 18 (10.5) | 0.98 (0.57-1.61) | 118 (10.4) | 5 (4.2) | 0.48 (0.17-1.10) | 54 (12.9) | 13 (24.1) | 1.58 (0.77-3.06) |
| Smoking or vaping | 37 (2.4) | 4 (10.8) | 1.02 (0.30-2.61) | 24 (2.1) | 2 (8.3) | 1.05 (0.17-3.63) | 13 (3.1) | 2 (15.4) | 0.84 (0.13-3.23) |
| Known COVID-19 exposure at home | 58 (3.7) | 12 (20.7) | 2.29 (1.14-4.29) | 28 (2.5) | 5 (17.9) | 2.59 (0.85-6.47) | 30 (7.2) | 7 (23.3) | 1.46 (0.56-3.39) |

<sup>1</sup> Odds ratios (OR) are unadjusted, comparing the selected group to the entire HCW population.
<sup>2</sup> OR for Other gender omitted due to small sample size.

Potential demographic risk factors included age, gender, race/ethnicity, and co-morbid conditions, as well as confirmed SARS-CoV-2 exposure outside the hospital (Table 1). No significant effect of age was noted among HCWs overall; however, a non-significant increase in seropositivity was observed for younger HCWs who were not previously tested by rt-PCR, indicating that younger HCW with SARS-CoV-2 infection may be less likely to screen positive for symptoms and be tested by rt-PCR. Male gender was associated with increased seropositivity, whereas race/ethnicity and co-morbidities were not. Confirmed COVID-19 exposure outside the hospital was the most significant demographic risk factor for seropositivity, with a higher OR among HCWs not tested by rt-PCR. These data indicate that either exposures occurred prior to widespread availability of rt-PCR testing which is unlikely given implementation of universal employee screening and testing early in the course of the local epidemic, or more likely that HCW were not always accurately reporting COVID-19 exposures during screening.

### Impact of Occupational Risk Factors on SARS-CoV-2 Seropositivity

Among roles within the hospital, only HCWs working in food services or environmental services showed significantly increased seropositivity as compared to the overall HCW population, and the effect was restricted to those not tested by rt-PCR. Similarly, working in administration was associated with increased seropositivity only amongst HCWs not tested by rt-PCR. Among locations in the hospital, working in COVID-19 units was associated with increased seropositivity, whereas working in labor and delivery units was associated with decreased seropositivity. COVID-19 patient contact and participation in aerosol-generating procedures on these patients were not associated with seropositivity.

Multivariable analyses included the non-occupational covariates discussed above, in addition to role and location within the hospital (Table 2). The Hosmer-Lemeshow goodness-of-fit test was non-significant (p=0.55) and area under the receiver-operating characteristic curve showed moderate discriminant ability (C-statistic=0.62), indicating that the adjusted model fit the data well.

**Table 2.** Associations between HCW occupational factors and SARS-CoV-2 seropositivity (AB prevalence) of HCW study population and subgroups segregated by prior rt-PCR testing.

| Variable | All HCWs (n=1,557) |  |  | Not tested by rt-PCR (n=1,138) |  |  | Tested by rt-PCR (n=419) |  |  |
| --- | --- | --- | --- | --- | --- | --- | --- | --- | --- |
|  | HCWs, n (%) | COVID-19 AB prevalence, n (%) | Adjusted OR (95% CI)<sup>1</sup> | HCWs, n (%) | COVID-19 AB prevalence, n (%) | Adjusted OR (95% CI)<sup>1</sup> | HCWs, n (%) | COVID-19 AB prevalence, n (%) | Adjusted OR (95% CI)<sup>1</sup> |
| Total | 1557 (100) | 165 (10.6) |  | 1138 (100) | 91 (8.0) |  | 419 (100) | 74 (17.7) |  |
| Role<sup>2</sup> |  |  |  |  |  |  |  |  |  |
| Physician | 246 (15.8) | 17 (6.9) | 0.59 (0.27-1.29) | 183 (16.1) | 10 (5.5) | 0.75 (0.27-2.15) | 63 (15.0) | 7 (11.1) | 0.41 (0.11-1.49) |
| Nurse | 705 (45.3) | 90 (12.8) | 1.47 (0.81-2.80) | 478 (42.0) | 40 (8.4) | 1.81 (0.81-4.55) | 227 (54.2) | 50 (22.0) | 1.24 (0.49-3.38) |
| Student | 69 (4.4) | 5 (7.2) | 0.75 (0.22-2.20) | 64 (5.6) | 5 (7.8) | 1.05 (0.28-3.59) | 5 (1.2) | 0 (0.0) | - |
| Ancillary clinical staff | 88 (5.7) | 7 (8.0) | 0.91 (0.32-2.36) | 55 (4.8) | 4 (7.3) | 1.48 (0.36-5.32) | 33 (7.9) | 3 (9.1) | 0.46 (0.08-2.19) |
| Administrative | 205 (13.2) | 23 (11.2) | 1.76 (0.86-3.69) | 164 (14.4) | 18 (11.0) | 2.69 (1.10-7.10) | 41 (9.8) | 5 (12.2) | 0.86 (0.21-3.30) |
| Food/environmental | 46 (3.0) | 7 (15.2) | 4.85 (1.51-14.85) | 37 (3.3) | 6 (16.2) | 8.28 (2.16-31.48) | 9 (2.1) | 1 (11.1) | 1.52 (0.06-17.75) |
| Other | 199 (12.8) | 16 (8.0) | 0.70 (0.39-1.18) | 157 (13.8) | 8 (5.1) | 0.54 (0.24-1.09) | 42 (10.0) | 8 (19.0) | 1.19 (0.48-2.65) |
| Location<sup>3</sup> |  |  |  |  |  |  |  |  |  |
| COVID ICU | 171 (11.0) | 26 (15.2) | 2.28 (1.29-3.96) | 100 (8.8) | 10 (10.0) | 2.35 (0.97-5.32) | 71 (16.9) | 16 (22.5) | 1.65 (0.71-3.79) |
| Non-COVID ICU | 364 (23.4) | 38 (10.4) | 0.89 (0.56-1.39) | 258 (22.7) | 18 (7.0) | 0.77 (0.40-1.40) | 106 (25.3) | 20 (18.9) | 1.04 (0.49-2.16) |
| COVID floor | 261 (16.8) | 35 (13.4) | 1.59 (1.01-2.48) | 133 (11.7) | 10 (7.5) | 1.21 (0.51-2.60) | 128 (30.5) | 25 (19.5) | 1.26 (0.67-2.35) |
| Non-COVID floor | 436 (28.0) | 50 (11.5) | 1.26 (0.85-1.85) | 309 (27.2) | 21 (6.8) | 0.85 (0.47-1.47) | 127 (30.3) | 29 (22.8) | 2.63 (1.40-4.97) |
| Labor and delivery | 113 (7.3) | 4 (3.5) | 0.24 (0.06-0.72) | 88 (7.7) | 4 (4.5) | 0.50 (0.12-1.56) | 25 (6.0) | 0 (0.0) | - |
| Operating room | 196 (12.6) | 15 (7.7) | 0.99 (0.53-1.78) | 173 (15.2) | 13 (7.5) | 1.13 (0.55-2.17) | 23 (5.5) | 2 (8.7) | 0.76 (0.11-3.43) |
| Non-operating room procedural | 198 (12.7) | 16 (8.1) | 0.91 (0.49-1.59) | 155 (13.6) | 14 (9.0) | 1.44 (0.72-2.74) | 43 (10.3) | 2 (4.7) | 0.36 (0.05-1.34) |
| Emergency department | 250 (16.1) | 20 (8.0) | 0.70 (0.40-1.18) | 199 (17.5) | 13 (6.5) | 0.62 (0.31-1.19) | 51 (12.2) | 7 (13.7) | 1.02 (0.36-2.67) |
| Outpatient clinical unit | 188 (12.1) | 13 (6.9) | 0.70 (0.36-1.27) | 143 (12.6) | 8 (5.6) | 0.72 (0.30-1.53) | 45 (10.7) | 5 (11.1) | 0.63 (0.19-1.73) |
| Non-clinical unit | 249 (16.0) | 21 (8.4) | 0.70 (0.37-1.24) | 201 (17.7) | 18 (9.0) | 0.76 (0.37-1.51) | 48 (11.5) | 3 (6.2) | 0.47 (0.09-1.76) |
| Job-related exposures<sup>4</sup> |  |  |  |  |  |  |  |  |  |
| Cared for COVID patient | 599 (38.5) | 69 (11.5) | 1.10 (0.79-1.54) | 396 (34.8) | 29 (7.3) | 0.85 (0.52-1.34) | 203 (48.4) | 40 (19.7) | 1.13 (0.67-1.92) |
| 3+ days in contact with COVID patient<sup>5</sup> | 263 (43.9) | 35 (13.3) | 1.39 (0.83-2.33) | 158 (39.9) | 19 (12.0) | 2.84 (1.27-6.69) | 105 (51.7) | 16 (15.2) | 0.61 (0.29-1.24) |
| Participated in aerosol-generating procedure<sup>5</sup> | 160 (26.7) | 15 (9.4) | 0.70 (0.37-1.27) | 116 (29.3) | 6 (5.2) | 0.51 (0.18-1.27) | 44 (21.7) | 9 (20.5) | 1.10 (0.44-2.55) |

<sup>1</sup> Adjusted ORs and 95% CI are adjusted for age, gender, race/ethnicity, known COVID exposure at home, role, location, and whether individual cared for COVID patient.
<sup>2</sup> Each role is compared to the entire HCW population, e.g., physicians vs. non-physicians.
<sup>3</sup> Individuals may select multiple locations, thus categories are not mutually exclusive. Each aOR corresponds to relative odds of being COVID AB-seropositive for individuals who worked in the specified location versus those who did not.
<sup>4</sup> HCWs may report multiple exposures or symptoms. Each aOR corresponds to relative odds of being COVID AB-seropositive for individuals who reported versus did not report the specified exposure or symptom.
<sup>5</sup> Days in contact with COVID patient and participated in aerosol-generating procedure only applicable for HCWs who reported “yes” to caring for COVID patients.

### Correlation of COVID-19 Symptoms with Seropositivity

A separate multivariable analysis was conducted that included non-occupational covariates discussed above, in addition to symptoms of COVID-19 (Table 3). Overall, multiple symptoms (fatigue, myalgias, fever, chills, and anosmia) were associated with increased seropositivity, with the strongest association observed for anosmia. These associations were entirely restricted to HCW tested previously by rt-PCR, indicating that occupational health screening was indeed effective in identifying symptomatic infections.

**Table 3.** Associations between HCW self-reported symptoms and SARS-CoV-2 seropositivity (AB prevalence) of HCW study population and subgroups segregated by prior rt-PCR testing.

| Variable | All HCWs (n=1,557) |  |  | Not tested by rt-PCR (n=1,138) |  |  | Tested by rt-PCR (n=419) |  |  |
| --- | --- | --- | --- | --- | --- | --- | --- | --- | --- |
|  | HCWs, n (%) | COVID-19 AB prevalence, n (%) | Adjusted OR (95% CI)<sup>1</sup> | HCWs, n (%) | COVID-19 AB prevalence, n (%) | Adjusted OR (95% CI)<sup>1</sup> | HCWs, n (%) | COVID-19 AB prevalence, n (%) | Adjusted OR (95% CI)<sup>1</sup> |
| Total | 1557 (100) | 165 (10.6) |  | 1138 (100) | 91 (8.0) |  | 419 (100) | 74 (17.7) |  |
| Symptoms<sup>2</sup> |  |  |  |  |  |  |  |  |  |
| Sore throat | 633 (40.7) | 79 (12.5) | 1.38 (1.00-1.92) | 405 (35.6) | 41 (10.1) | 1.47 (0.95-2.27) | 228 (54.4) | 38 (16.7) | 0.95 (0.56-1.60) |
| Fatigue | 429 (27.6) | 63 (14.7) | 1.77 (1.25-2.49) | 244 (21.4) | 17 (7.0) | 0.79 (0.44-1.35) | 185 (44.2) | 46 (24.9) | 2.94 (1.72-5.12) |
| Muscle aches | 361 (23.2) | 55 (15.2) | 1.76 (1.23-2.50) | 198 (17.4) | 15 (7.6) | 0.89 (0.48-1.56) | 163 (38.9) | 40 (24.5) | 2.20 (1.30-3.74) |
| New cough | 470 (30.2) | 54 (11.5) | 1.11 (0.78-1.57) | 292 (25.7) | 18 (6.2) | 0.65 (0.36-1.09) | 178 (42.5) | 36 (20.2) | 1.50 (0.89-2.53) |
| New chills | 327 (21.0) | 51 (15.6) | 1.79 (1.24-2.55) | 183 (16.1) | 12 (6.6) | 0.74 (0.37-1.34) | 144 (34.4) | 39 (27.1) | 2.61 (1.55-4.44) |
| Fever | 318 (20.4) | 48 (15.1) | 1.67 (1.15-2.39) | 177 (15.6) | 11 (6.2) | 0.67 (0.33-1.24) | 141 (33.7) | 37 (26.2) | 2.38 (1.40-4.04) |
| Anosmia | 95 (6.1) | 33 (34.7) | 5.34 (3.33-8.45) | 40 (3.5) | 5 (12.5) | 1.67 (0.56-4.08) | 55 (13.1) | 28 (50.9) | 7.67 (4.05-14.76) |
| Dyspnea | 200 (12.8) | 27 (13.5) | 1.38 (0.87-2.13) | 113 (9.9) | 8 (7.1) | 0.83 (0.36-1.68) | 87 (20.8) | 19 (21.8) | 1.61 (0.86-2.91) |

<sup>1</sup> Adjusted ORs and 95% CI are adjusted for age, gender, race/ethnicity, known COVID exposure at home, role, location, and whether individual cared for COVID-19 patient.
<sup>2</sup> HCWs may report multiple exposures and/or symptoms. Each adjusted OR corresponds to relative odds of being COVID AB seropositive for individuals who reported versus did not report the specified exposure or symptom.

## Discussion

This study provides several insights into the relationships between non-occupational and occupational risk factors and COVID-19 seropositivity among HCWs (Figure 1). Exposure to COVID-19 outside of work was a greater risk factor for seropositivity than any occupational exposure other than working in food or environmental services. The HCW roles associated with the greatest risk of seropositivity did not involve direct patient care. Nurses, who have the most direct and sustained patient contact, were not at significantly increased risk. The only locations associated with increased seropositivity were the dedicated COVID-19 ICU and floor units. The operating room, an area of great concern due to intubation of multiple patients, was not associated with increased risk. Performing aerosol-generating procedures on known COVID-19 patients was also not significantly associated with seropositivity, which is reassuring given that perceived risk of transmission during these procedures can delay patient care.

**Figure 1.**
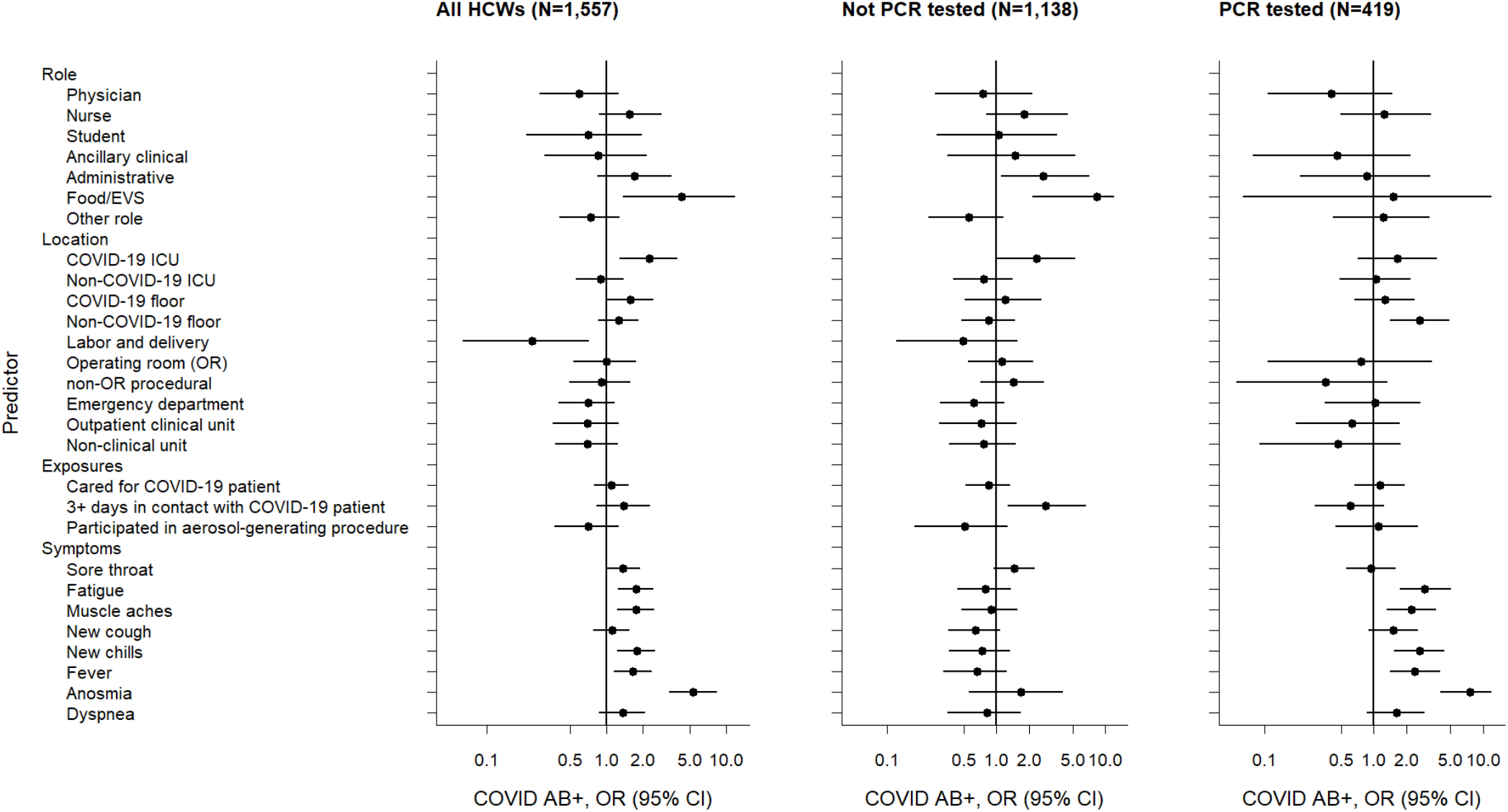
Forest plot of adjusted odds ratios (OR) of hypothesized predictors of COVID-19 seropositivity (AB+) among HCW study population and subgroups segregated by prior rt-PCR testing. ORs are adjusted for sex, age, race/ethnicity, known COVID-19 exposure outside of work, role, location, and COVID-19 patient contact. (EVS, environmental services)

Stratification of HCWs based on whether or not they were tested previously by rt-PCR yielded several additional insights into the strengths and weaknesses of universal symptom screening. The association between COVID-19 symptoms and seropositivity was restricted to HCW tested previously by rt-PCR, indicating that universal screening was effective in identifying symptomatic infections. Younger HCWs who were COVID-19-seropositive were more likely to be missed by occupational screening, which is consistent with the increased prevalence of minimally symptomatic infection among younger individuals[27]. Decreased seropositivity among HCW in labor and delivery units may be due to increased vigilance amongst HCW who care for pregnant patients or low disease prevalence among these patients. While the hospital’s mandatory screening was only implemented one month prior to this study, the prevalence of COVID-19 in Orange County was low at that time and increased subsequently (Figure 2). The CoVAM was trained and validated on blood specimens collected relatively soon after symptom onset (minimum 7 d, median 11 d) consistent with known kinetics of the antibody response to COVID-19 [3]. Therefore, we believe the effects of occupational health screening are at least partially captured in our analysis. In addition, the study hospital was able to maintain infection prevention best practices consistent with guidance from the U.S. Centers for Disease Control and Prevention (CDC), including continuous, ample availability of PPE throughout the pandemic.

**Figure 2.**
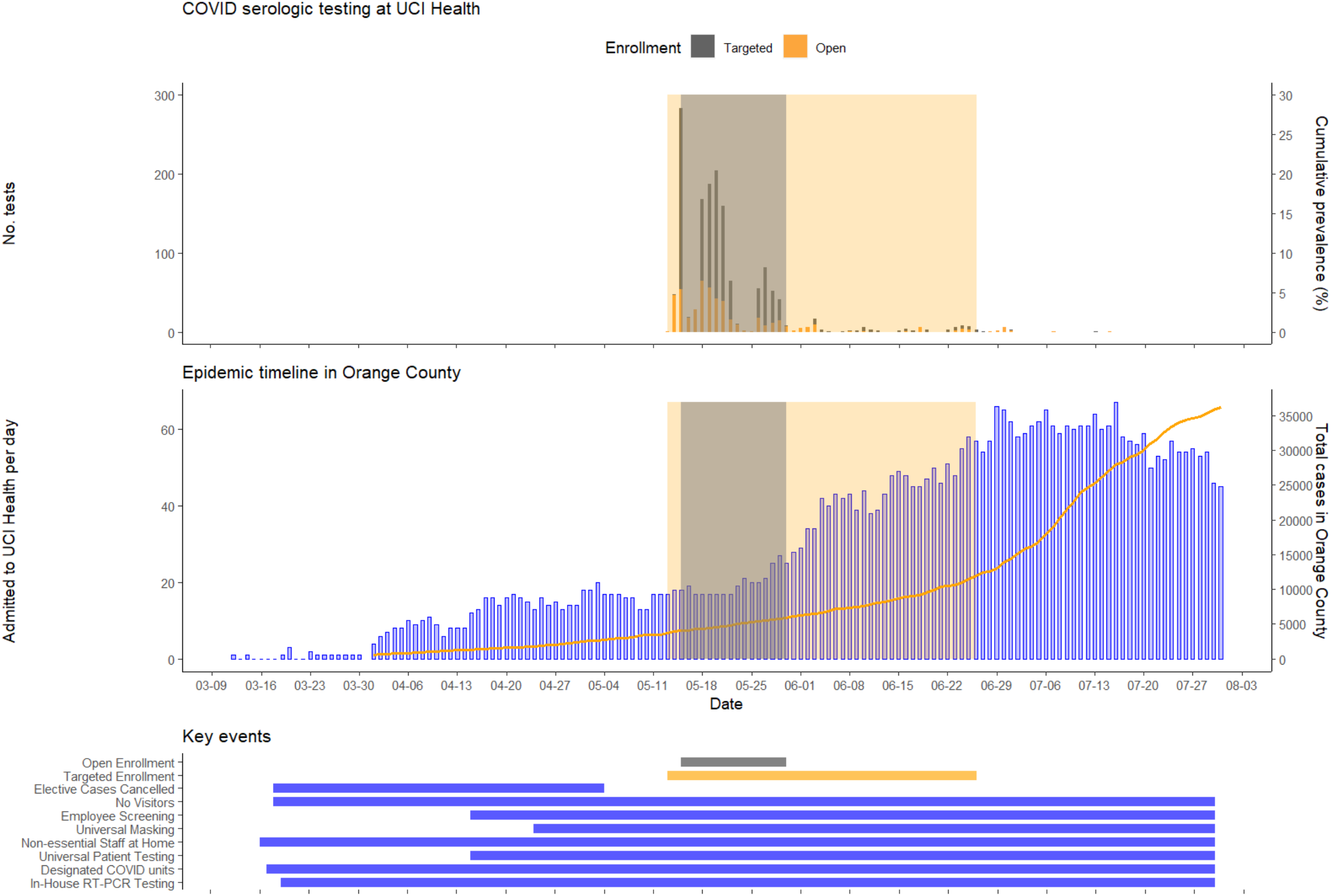
Epidemiologic context of HCW study with respect to community prevalence and hospital burden of COVID-19.

No data were available for SARS-CoV-2 seropositivity in the surrounding community at the time that the study was performed. The cumulative COVID-19 prevalence in Orange County at the time of this study’s enrollment was 0.2%. The overall seropositivity rate was 10.8% in this study, but the 8.0% seropositivity amongst HCW not previously identified by screening, which matches the seropositivity in the open enrollment cohort, is most appropriate for comparison to community prevalence to avoid the enrichment effect of the targeted enrollment cohort. This seropositivity is 40-fold higher than community prevalence as determined by rt-PCR testing (Figure 2). Whereas more recent seroprevalence studies show a 10-fold increase compared to case counts, our result is most comparable to early seroprevalence studies prior to significant local outbreaks of COVID-19 that have found larger disparities between seropositivity rates and case counts[5-9, 13]. Recently, a community study sampled 2,979 random participants in Orange County from July 10 through August 16, 2020 and found a seropositivity rate of 11.5% using an updated version of the CoVAM with a more stringent threshold for seropositivity[28]. Taken together, these observations imply that HCW are not more likely to be seropositive than the surrounding community, although the different study periods limit this direct comparison.

The findings of this study are largely consistent with recently published studies of COVID-19 seropositivity among HCWs[18, 19]. In particular, the lack of association of either race/ethnicity or co-morbidities with seropositivity in our study is consistent with these prior HCW studies and differs from a prior community study that did observe such associations[29]. This study provides additional insight compared to prior studies by examining specific roles in the hospital and controlling for multiple likely sources of confounding. For example, nurses had significantly elevated seropositivity in bivariate analyses (unadjusted OR [CI]=1.52 [1.10-2.10]) but this finding did not persist after adjusting for work location; in contrast, null associations between seropositivity and roles without direct patient care became significant and positive after adjusting for location (unadjusted OR [CI] for administrative = 1.08 [0.66, 1.69]; food/environmental = 1.54 [0.62-3.29]).

Strengths of this study include the validated test performance of the CoVAM, which compares favorably to currently available single-antigen assays; the large sample size with inclusion of 34.4% of HCW at the hospital; and the use of multivariable analysis to control for confounding. The weaknesses of this study include the non-random enrollment methodology as the targeted enrollment cohort was invited from groups expected to have higher seroprevalence and the open enrollment cohort was subject to self-selection, which both could lead to sampling bias. Also, different blood sampling methodology was used in the open and targeted enrollment cohorts due to institutional interest in banking specimens from the latter group. The subgroup analysis based on prior rt-PCR testing was used to control for the heterogeneous sampling, as prior testing was the primary driver of increased prevalence in the targeted enrollment cohort. When the study population is stratified based on method of recruitment (Supplementary Tables 1-3), the results are largely similar to stratification based on rt-PCR testing (Tables 1-3).

The results of this study have several implications for the local and global responses to the COVID-19 pandemic. The finding of a significantly increased SARS-CoV-2 prevalence by serology as compared with rt-PCR provides evidence that the reported counts of confirmed COVID-19 cases are significant underestimates. The observations that HCWs who are younger, work in non-patient care roles, or have COVID-19 exposure outside of work are more likely to have COVID-19 seropositivity without prior testing indicates that screening and vaccination efforts need to focus on these groups. The lack of association of aerosol-generating procedures with COVID-19 seropositivity in the context of adequate availability and presumably appropriate use of PPE is reassuring. The increased seropositivity in COVID-19 units is less reassuring but may potentially be explained by employee-to-employee spread in these locations given that caring for COVID-19 patients was not a significant risk factor. Further studies that include longitudinal follow-up are needed to confirm these observations.

## Data Availability

The dataset generated by testing specimens on the coronavirus antigen microarray and the analysis code applied to this dataset is available upon request. The associated clinical data with removal of all identifying information is also available upon request.

## Author Contributions

The study was designed by S. Schubl and S. Khan and coordinated by C. Figueroa, and study procedures were conducted by D. Brabender, A. Naaseh, O. Dominguez, A. Runge, S. Skochko, and J. Chinn. Blood specimens were processed by P. Horvath under the supervision of F. Zaldivar and by D. Tifrea under the supervision of R. Edwards. The coronavirus antigen microarray was designed by S. Khan and P. Felgner and was constructed by R. Nakajima. The testing of specimens on the coronavirus antigen microarray was performed by A. Jain under the supervision of S. Khan and P. Felgner and A. Kelsey and K. Lai under the supervision of W. Zhao. The CoVAM data was analyzed by R. de Assis under the supervision of S. Khan and P. Felgner. The study data was collated by C. Figueroa and A. Gonzales and analyzed by A. Palma and A. Grigorian. The data was interpreted and the manuscript and figures were prepared by S. Khan, S. Schubl, and A. Palma with substantial input from M. Stamos, A. Amin and P. Barie and additional input and approval from all other authors.

## Acknowledgements

This work was supported by two intramural research grants from the COVID-19 Basic, Translational, and Clinical Research Fund of the University of California Irvine and by the Emergency COVID-19 Research Seed Funding Opportunity from the University of California Office of the President [research grants R00RG2646, R01RG3745]. The findings and conclusions in this report are those of the authors and do not necessarily represent the official position or policy of the University of California.

This work utilized the Chao Family Comprehensive Cancer Center Experimental Tissue Shared Resource, supported by the National Cancer Institute of the National Institutes of Health [award number P30CA062203].

S. Khan was supported by the National Center for Research Resources and the National Center for Advancing Translational Sciences of the National Institutes of Health [grant KL2 TR001416]. The project described was supported by the National Center for Research Resources and the National Center for Advancing Translational Sciences of the National Institutes of Health [grant UL1 TR001414]. The content is solely the responsibility of the authors and does not necessarily represent the official views of the NIH.

The initial design and construction of the CoVAM was supported by the Prometheus-UMD contract sponsored by the Defense Advanced Research Projects Agency (DARPA) BTO under the auspices of Col. Matthew Hepburn [agreements N66001-17-2-4023, N66001-18-2-4015]. The findings and conclusions in this report are those of the authors and do not necessarily represent the official position or policy of the funding agencies and no official endorsements should be inferred.

## Materials and Correspondence

Please address all correspondence and material requests related to this manuscript to Saahir Khan. The dataset generated by testing specimens on the coronavirus antigen microarray and the analysis code applied to this dataset is available upon request. The associated clinical data with removal of all identifying information is also available upon request.

## Supplementary Information

### Supplementary Tables

**Supplementary Table 1.**
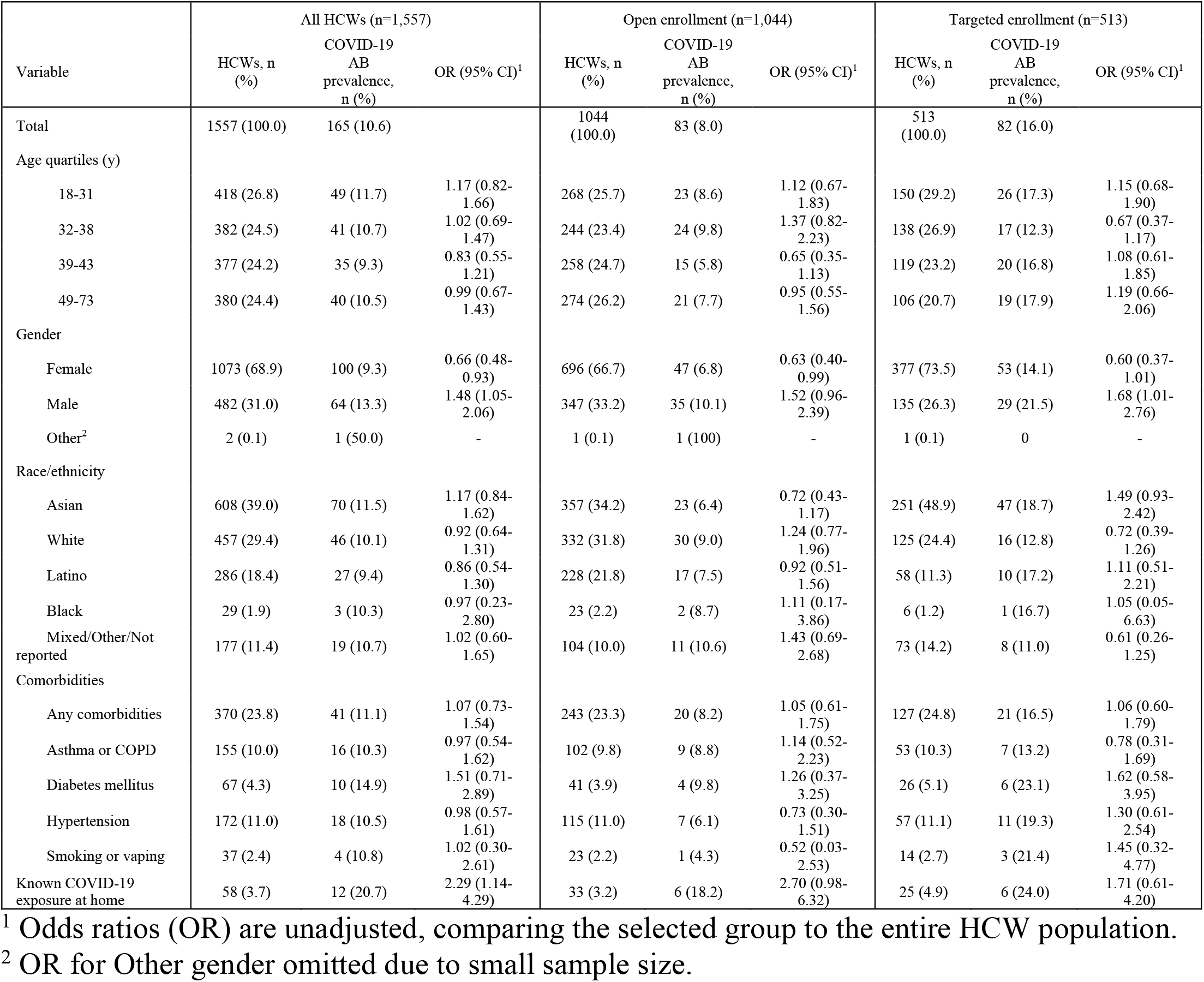
Association between demographic and health-related characteristics and SARS-CoV-2 seropositivity (AB prevalence) of HCW study population and subgroups segregated by enrollment group. (COPD, chronic obstructive pulmonary disease)

| Variable | All HCWs (n=1,557) |  |  | Open enrollment (n=1,044) |  |  | Targeted enrollment (n=513) |  |  |
| --- | --- | --- | --- | --- | --- | --- | --- | --- | --- |
|  | HCWs, n (%) | COVID-19 AB prevalence, n (%) | OR (95% CI)<sup>1</sup> | HCWs, n (%) | COVID-19 AB prevalence, n (%) | OR (95% CI)<sup>1</sup> | HCWs, n (%) | COVID-19 AB prevalence, n (%) | OR (95% CI)<sup>1</sup> |
| Total | 1557 (100.0) | 165 (10.6) |  | 1044 (100.0) | 83 (8.0) |  | 513 (100.0) | 82 (16.0) |  |
| Age quartiles (y) |  |  |  |  |  |  |  |  |  |
| 18-31 | 418 (26.8) | 49 (11.7) | 1.17 (0.82-1.66) | 268 (25.7) | 23 (8.6) | 1.12 (0.67-1.83) | 150 (29.2) | 26 (17.3) | 1.15 (0.68-1.90) |
| 32-38 | 382 (24.5) | 41 (10.7) | 1.02 (0.69-1.47) | 244 (23.4) | 24 (9.8) | 1.37 (0.82-2.23) | 138 (26.9) | 17 (12.3) | 0.67 (0.37-1.17) |
| 39-43 | 377 (24.2) | 35 (9.3) | 0.83 (0.55-1.21) | 258 (24.7) | 15 (5.8) | 0.65 (0.35-1.13) | 119 (23.2) | 20 (16.8) | 1.08 (0.61-1.85) |
| 49-73 | 380 (24.4) | 40 (10.5) | 0.99 (0.67-1.43) | 274 (26.2) | 21 (7.7) | 0.95 (0.55-1.56) | 106 (20.7) | 19 (17.9) | 1.19 (0.66-2.06) |
| Gender |  |  |  |  |  |  |  |  |  |
| Female | 1073 (68.9) | 100 (9.3) | 0.66 (0.48-0.93) | 696 (66.7) | 47 (6.8) | 0.63 (0.40-0.99) | 377 (73.5) | 53 (14.1) | 0.60 (0.37-1.01) |
| Male | 482 (31.0) | 64 (13.3) | 1.48 (1.05-2.06) | 347 (33.2) | 35 (10.1) | 1.52 (0.96-2.39) | 135 (26.3) | 29 (21.5) | 1.68 (1.01-2.76) |
| Other<sup>2</sup> | 2 (0.1) | 1 (50.0) | - | 1 (0.1) | 1 (100) | - | 1 (0.1) | 0 | - |
| Race/ethnicity |  |  |  |  |  |  |  |  |  |
| Asian | 608 (39.0) | 70 (11.5) | 1.17 (0.84-1.62) | 357 (34.2) | 23 (6.4) | 0.72 (0.43-1.17) | 251 (48.9) | 47 (18.7) | 1.49 (0.93-2.42) |
| White | 457 (29.4) | 46 (10.1) | 0.92 (0.64-1.31) | 332 (31.8) | 30 (9.0) | 1.24 (0.77-1.96) | 125 (24.4) | 16 (12.8) | 0.72 (0.39-1.26) |
| Latino | 286 (18.4) | 27 (9.4) | 0.86 (0.54-1.30) | 228 (21.8) | 17 (7.5) | 0.92 (0.51-1.56) | 58 (11.3) | 10 (17.2) | 1.11 (0.51-2.21) |
| Black | 29 (1.9) | 3 (10.3) | 0.97 (0.23-2.80) | 23 (2.2) | 2 (8.7) | 1.11 (0.17-3.86) | 6 (1.2) | 1 (16.7) | 1.05 (0.05-6.63) |
| Mixed/Other/Not reported | 177 (11.4) | 19 (10.7) | 1.02 (0.60-1.65) | 104 (10.0) | 11 (10.6) | 1.43 (0.69-2.68) | 73 (14.2) | 8 (11.0) | 0.61 (0.26-1.25) |
| Comorbidities |  |  |  |  |  |  |  |  |  |
| Any comorbidities | 370 (23.8) | 41 (11.1) | 1.07 (0.73-1.54) | 243 (23.3) | 20 (8.2) | 1.05 (0.61-1.75) | 127 (24.8) | 21 (16.5) | 1.06 (0.60-1.79) |
| Asthma or COPD | 155 (10.0) | 16 (10.3) | 0.97 (0.54-1.62) | 102 (9.8) | 9 (8.8) | 1.14 (0.52-2.23) | 53 (10.3) | 7 (13.2) | 0.78 (0.31-1.69) |
| Diabetes mellitus | 67 (4.3) | 10 (14.9) | 1.51 (0.71-2.89) | 41 (3.9) | 4 (9.8) | 1.26 (0.37-3.25) | 26 (5.1) | 6 (23.1) | 1.62 (0.58-3.95) |
| Hypertension | 172 (11.0) | 18 (10.5) | 0.98 (0.57-1.61) | 115 (11.0) | 7 (6.1) | 0.73 (0.30-1.51) | 57 (11.1) | 11 (19.3) | 1.30 (0.61-2.54) |
| Smoking or vaping | 37 (2.4) | 4 (10.8) | 1.02 (0.30-2.61) | 23 (2.2) | 1 (4.3) | 0.52 (0.03-2.53) | 14 (2.7) | 3 (21.4) | 1.45 (0.32-4.77) |
| Known COVID-19 exposure at home | 58 (3.7) | 12 (20.7) | 2.29 (1.14-4.29) | 33 (3.2) | 6 (18.2) | 2.70 (0.98-6.32) | 25 (4.9) | 6 (24.0) | 1.71 (0.61-4.20) |

<sup>1</sup> Odds ratios (OR) are unadjusted, comparing the selected group to the entire HCW population.
<sup>2</sup> OR for Other gender omitted due to small sample size.

**Supplementary Table 2.**
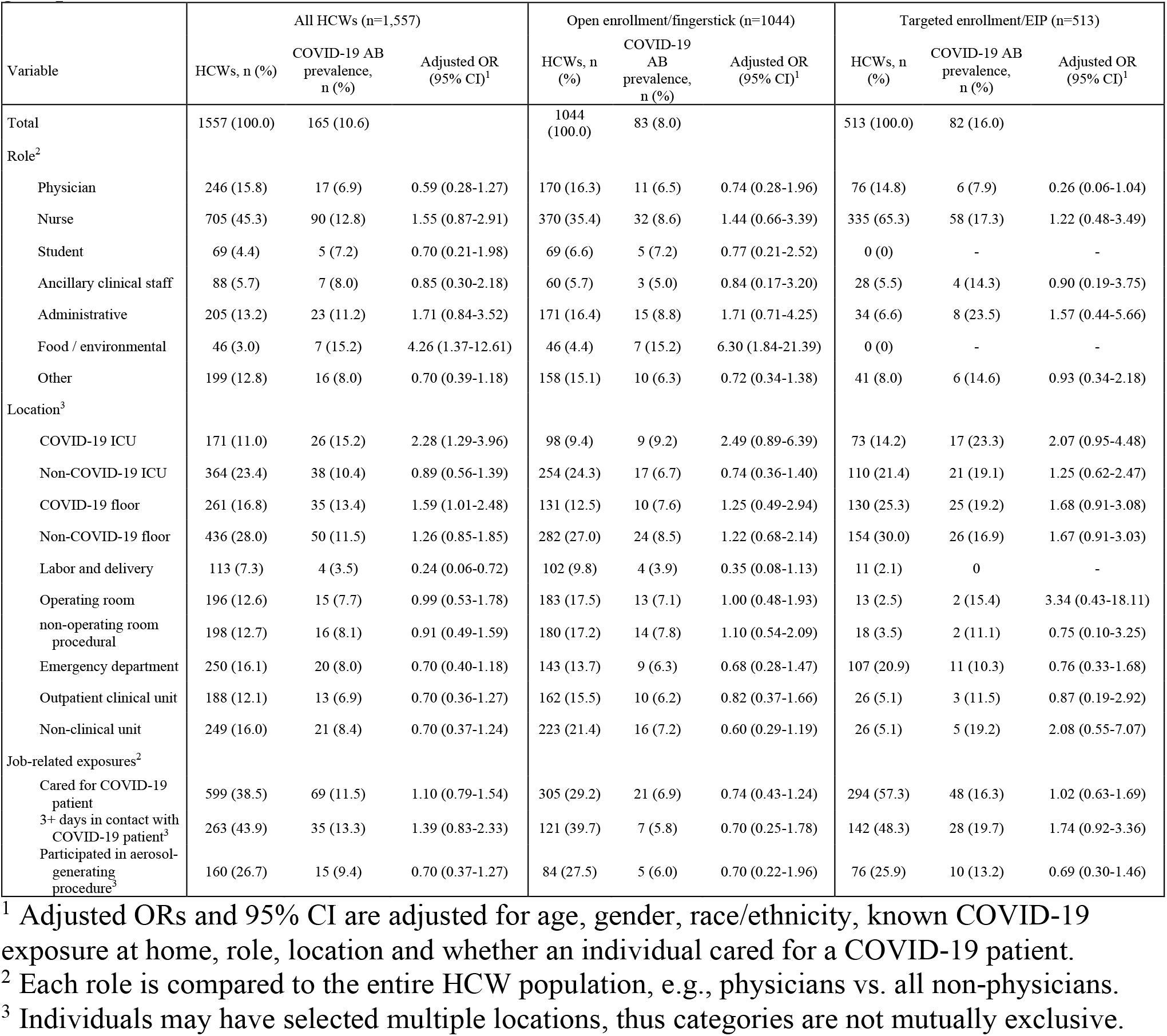
Association between HCW occupational factors and SARS-CoV-2 seropositivity (AB prevalence) of HCW study population and subgroups segregated by enrollment group.

| Variable | All HCWs (n=1,557) |  |  | Open enrollment/fingerstick (n=1044) |  |  | Targeted enrollment/EIP (n=513) |  |  |
| --- | --- | --- | --- | --- | --- | --- | --- | --- | --- |
|  | HCWs, n (%) | COVID-19 AB prevalence, n (%) | Adjusted OR (95% CI)<sup>1</sup> | HCWs, n (%) | COVID-19 AB prevalence, n (%) | Adjusted OR (95% CI)<sup>1</sup> | HCWs, n (%) | COVID-19 AB prevalence, n (%) | Adjusted OR (95% CI)<sup>1</sup> |
| Total | 1557 (100.0) | 165 (10.6) |  | 1044 (100.0) | 83 (8.0) |  | 513 (100.0) | 82 (16.0) |  |
| Role<sup>2</sup> |  |  |  |  |  |  |  |  |  |
| Physician | 246 (15.8) | 17 (6.9) | 0.59 (0.28-1.27) | 170 (16.3) | 11 (6.5) | 0.74 (0.28-1.96) | 76 (14.8) | 6 (7.9) | 0.26 (0.06-1.04) |
| Nurse | 705 (45.3) | 90 (12.8) | 1.55 (0.87-2.91) | 370 (35.4) | 32 (8.6) | 1.44 (0.66-3.39) | 335 (65.3) | 58 (17.3) | 1.22 (0.48-3.49) |
| Student | 69 (4.4) | 5 (7.2) | 0.70 (0.21-1.98) | 69 (6.6) | 5 (7.2) | 0.77 (0.21-2.52) | 0 (0) | - | - |
| Ancillary clinical staff | 88 (5.7) | 7 (8.0) | 0.85 (0.30-2.18) | 60 (5.7) | 3 (5.0) | 0.84 (0.17-3.20) | 28 (5.5) | 4 (14.3) | 0.90 (0.19-3.75) |
| Administrative | 205 (13.2) | 23 (11.2) | 1.71 (0.84-3.52) | 171 (16.4) | 15 (8.8) | 1.71 (0.71-4.25) | 34 (6.6) | 8 (23.5) | 1.57 (0.44-5.66) |
| Food / environmental | 46 (3.0) | 7 (15.2) | 4.26 (1.37-12.61) | 46 (4.4) | 7 (15.2) | 6.30 (1.84-21.39) | 0 (0) | - | - |
| Other | 199 (12.8) | 16 (8.0) | 0.70 (0.39-1.18) | 158 (15.1) | 10 (6.3) | 0.72 (0.34-1.38) | 41 (8.0) | 6 (14.6) | 0.93 (0.34-2.18) |
| Location<sup>3</sup> |  |  |  |  |  |  |  |  |  |
| COVID-19 ICU | 171 (11.0) | 26 (15.2) | 2.28 (1.29-3.96) | 98 (9.4) | 9 (9.2) | 2.49 (0.89-6.39) | 73 (14.2) | 17 (23.3) | 2.07 (0.95-4.48) |
| Non-COVID-19 ICU | 364 (23.4) | 38 (10.4) | 0.89 (0.56-1.39) | 254 (24.3) | 17 (6.7) | 0.74 (0.36-1.40) | 110 (21.4) | 21 (19.1) | 1.25 (0.62-2.47) |
| COVID-19 floor | 261 (16.8) | 35 (13.4) | 1.59 (1.01-2.48) | 131 (12.5) | 10 (7.6) | 1.25 (0.49-2.94) | 130 (25.3) | 25 (19.2) | 1.68 (0.91-3.08) |
| Non-COVID-19 floor | 436 (28.0) | 50 (11.5) | 1.26 (0.85-1.85) | 282 (27.0) | 24 (8.5) | 1.22 (0.68-2.14) | 154 (30.0) | 26 (16.9) | 1.67 (0.91-3.03) |
| Labor and delivery | 113 (7.3) | 4 (3.5) | 0.24 (0.06-0.72) | 102 (9.8) | 4 (3.9) | 0.35 (0.08-1.13) | 11 (2.1) | 0 | - |
| Operating room | 196 (12.6) | 15 (7.7) | 0.99 (0.53-1.78) | 183 (17.5) | 13 (7.1) | 1.00 (0.48-1.93) | 13 (2.5) | 2 (15.4) | 3.34 (0.43-18.11) |
| non-operating room procedural | 198 (12.7) | 16 (8.1) | 0.91 (0.49-1.59) | 180 (17.2) | 14 (7.8) | 1.10 (0.54-2.09) | 18 (3.5) | 2 (11.1) | 0.75 (0.10-3.25) |
| Emergency department | 250 (16.1) | 20 (8.0) | 0.70 (0.40-1.18) | 143 (13.7) | 9 (6.3) | 0.68 (0.28-1.47) | 107 (20.9) | 11 (10.3) | 0.76 (0.33-1.68) |
| Outpatient clinical unit | 188 (12.1) | 13 (6.9) | 0.70 (0.36-1.27) | 162 (15.5) | 10 (6.2) | 0.82 (0.37-1.66) | 26 (5.1) | 3 (11.5) | 0.87 (0.19-2.92) |
| Non-clinical unit | 249 (16.0) | 21 (8.4) | 0.70 (0.37-1.24) | 223 (21.4) | 16 (7.2) | 0.60 (0.29-1.19) | 26 (5.1) | 5 (19.2) | 2.08 (0.55-7.07) |
| Job-related exposures<sup>2</sup> |  |  |  |  |  |  |  |  |  |
| Cared for COVID-19 patient | 599 (38.5) | 69 (11.5) | 1.10 (0.79-1.54) | 305 (29.2) | 21 (6.9) | 0.74 (0.43-1.24) | 294 (57.3) | 48 (16.3) | 1.02 (0.63-1.69) |
| 3+ days in contact with COVID-19 patient<sup>3</sup> | 263 (43.9) | 35 (13.3) | 1.39 (0.83-2.33) | 121 (39.7) | 7 (5.8) | 0.70 (0.25-1.78) | 142 (48.3) | 28 (19.7) | 1.74 (0.92-3.36) |
| Participated in aerosol-generating procedure<sup>3</sup> | 160 (26.7) | 15 (9.4) | 0.70 (0.37-1.27) | 84 (27.5) | 5 (6.0) | 0.70 (0.22-1.96) | 76 (25.9) | 10 (13.2) | 0.69 (0.30-1.46) |

<sup>1</sup> Adjusted ORs and 95% CI are adjusted for age, gender, race/ethnicity, known COVID-19 exposure at home, role, location and whether an individual cared for a COVID-19 patient.
<sup>2</sup> Each role is compared to the entire HCW population, e.g., physicians vs. all non-physicians.
<sup>3</sup> Individuals may have selected multiple locations, thus categories are not mutually exclusive.

**Supplementary Table 3.** Association between HCW self-reported symptoms and SARS-CoV-2 seropositivity (AB prevalence) of HCW study population and subgroups segregated by enrollment group.

| Variable | All HCWs (n=1,557) |  |  | Open enrollment (n=1,044) |  |  | Targeted enrollment (n=513) |  |  |
| --- | --- | --- | --- | --- | --- | --- | --- | --- | --- |
|  | HCWs, n (%) | COVID-19 AB prevalence, n (%) | Adjusted OR (95% CI)<sup>1</sup> | HCWs, n (%) | COVID-19 AB prevalence, n (%) | Adjusted OR (95% CI)<sup>1</sup> | HCWs, n (%) | COVID-19 AB prevalence, n (%) | Adjusted OR (95% CI)<sup>1</sup> |
| Total | 1557 (100.0) | 165 (10.6) |  | 1044 (100.0) | 83 (8.0) |  | 513 (100.0) | 82 (16.0) |  |
| Symptoms<sup>2</sup> |  |  |  |  |  |  |  |  |  |
| Sore throat | 633 (40.7) | 79 (12.5) | 1.38 (1.00-1.92) | 415 (39.8) | 35 (8.4) | 1.07 (0.67-1.70) | 218 (42.5) | 44 (20.2) | 1.72 (1.06-2.79) |
| Fatigue | 429 (27.6) | 63 (14.7) | 1.77 (1.25-2.49) | 259 (24.8) | 20 (7.7) | 0.91 (0.52-1.54) | 170 (33.1) | 43 (25.3) | 2.75 (1.68-4.52) |
| Muscle aches | 361 (23.2) | 55 (15.2) | 1.76 (1.23-2.50) | 216 (20.7) | 15 (6.9) | 0.77 (0.41-1.35) | 145 (28.3) | 40 (27.6) | 2.95 (1.79-4.88) |
| New cough | 470 (30.2) | 54 (11.5) | 1.11 (0.78-1.57) | 305 (29.2) | 22 (7.2) | 0.81 (0.47-1.33) | 165 (32.2) | 32 (19.4) | 1.41 (0.85-2.31) |
| New chills | 327 (21.0) | 51 (15.6) | 1.79 (1.24-2.55) | 203 (19.4) | 17 (8.4) | 0.99 (0.54-1.70) | 124 (24.2) | 34 (27.4) | 2.59 (1.55-4.29) |
| Fever | 318 (20.4) | 48 (15.1) | 1.67 (1.15-2.39) | 191 (18.3) | 15 (7.9) | 0.90 (0.48-1.59) | 127 (24.8) | 33 (26.0) | 2.34 (1.40-3.86) |
| Loss of smell | 95 (6.1) | 33 (34.7) | 5.34 (3.33-8.45) | 51 (4.9) | 9 (17.6) | 2.47 (1.07-5.12) | 44 (8.6) | 24 (54.5) | 8.50 (4.37-16.75) |
| Dyspnea | 200 (12.8) | 27 (13.5) | 1.38 (0.87-2.13) | 131 (12.5) | 12 (9.2) | 1.11 (0.54-2.08) | 69 (13.5) | 15 (21.7) | 1.49 (0.77-2.77) |

<sup>1</sup> Adjusted ORs and 95% CI are adjusted for age, gender, race/ethnicity, known COVID-19 exposure at home, role, location and whether an individual cared for a COVID-19 patient.
<sup>2</sup> HCWs may have reported multiple exposures or symptoms. Each adjusted OR corresponds to relative odds of being COVID-19 AB-seropositive for individuals who reported versus did not report the specified exposure or symptom.
<sup>3</sup> Days in contact with a COVID-19 patient and participated in aerosol-generating procedure only applicable for HCWs who reported “yes” to caring for COVID-19 patients

**Supplementary Table 4.** Demographic and health-related characteristics and SARS-CoV-2 seropositivity of HCW subgroups without prior rt-PCR testing and with prior positive and negative rt-PCR testing.

| Variable | Not tested for PCR |  | PCR+ |  | PCR- |  |
| --- | --- | --- | --- | --- | --- | --- |
|  | HCWs, n (%) | COVID AB+, n (%) | HCWs, n (%) | COVID AB+, n (%) | HCWs, n (%) | COVID AB+, n (%) |
| **Total** | 1138 (100.0) | 91 (8.0) | 38 (100.0) | 36 (94.7) | 322 (100.0) | 30 (9.3) |
| **Age quartiles (y)** |  |  |  |  |  |  |
| 18-31 | 311 (27.3) | 33 (10.6) | 7 (18.4) | 6 (85.7) | 86 (26.7) | 7 (8.1) |
| 32-38 | 257 (22.6) | 22 (8.6) | 9 (23.7) | 8 (88.9) | 98 (30.4) | 10 (10.2) |
| 39-43 | 275 (24.2) | 15 (5.5) | 11 (28.9) | 11 (100.0) | 74 (23.0) | 7 (9.5) |
| 49-73 | 295 (25.9) | 21 (7.1) | 11 (28.9) | 11 (100.0) | 64 (19.9) | 6 (9.4) |
| **Gender** |  |  |  |  |  |  |
| Female | 781 (68.6) | 57 (7.3) | 22 (57.9) | 21 (95.5) | 228 (70.8) | 18 (7.9) |
| Male | 355 (31.2) | 33 (9.3) | 16 (42.1) | 15 (93.8) | 94 (29.2) | 12 (12.8) |
| **Race/ethnicity** |  |  |  |  |  |  |
| Asian | 415 (36.5) | 26 (6.3) | 23 (60.5) | 22 (95.7) | 151 (46.9) | 19 (12.6) |
| White | 336 (29.5) | 32 (9.5) | 6 (15.8) | 6 (100.0) | 92 (28.6) | 6 (6.5) |
| Latino | 232 (20.4) | 18 (7.8) | 7 (18.4) | 7 (100.0) | 45 (14.0) | 1 (2.2) |
| Black | 25 (2.2) | 2 (8.0) | NA | NA | 3 (0.9) | 1 (33.3) |
| Mixed/Other/Not reported | 130 (11.4) | 13 (10.0) | 2 (5.3) | 1 (50.0) | 31 (9.6) | 3 (9.7) |
| **Comorbidities** |  |  |  |  |  |  |
| Any comorbidities | 258 (22.7) | 18 (7.0) | 14 (36.8) | 14 (100.0) | 80 (24.8) | 7 (8.8) |
| Asthma or COPD | 114 (10.0) | 9 (7.9) | 3 (7.9) | 3 (100.0) | 31 (9.6) | 2 (6.5) |
| Diabetes mellitus | 45 (4.0) | 3 (6.7) | 4 (10.5) | 4 (100.0) | 14 (4.3) | 3 (21.4) |
| Hypertension | 118 (10.4) | 5 (4.2) | 9 (23.7) | 9 (100.0) | 39 (12.1) | 4 (10.3) |
| Smoking or vaping | 24 (2.1) | 2 (8.3) | 1 (2.6) | 1 (100.0) | 10 (3.1) | 1 (10.0) |
| **Known COVID-19 exposure at home** | 28 (2.5) | 5 (17.9) | 5 (13.2) | 5 (100.0) | 24 (7.5) | 2 (8.3) |

### Supplementary Figures

**Supplementary Figure 1.**
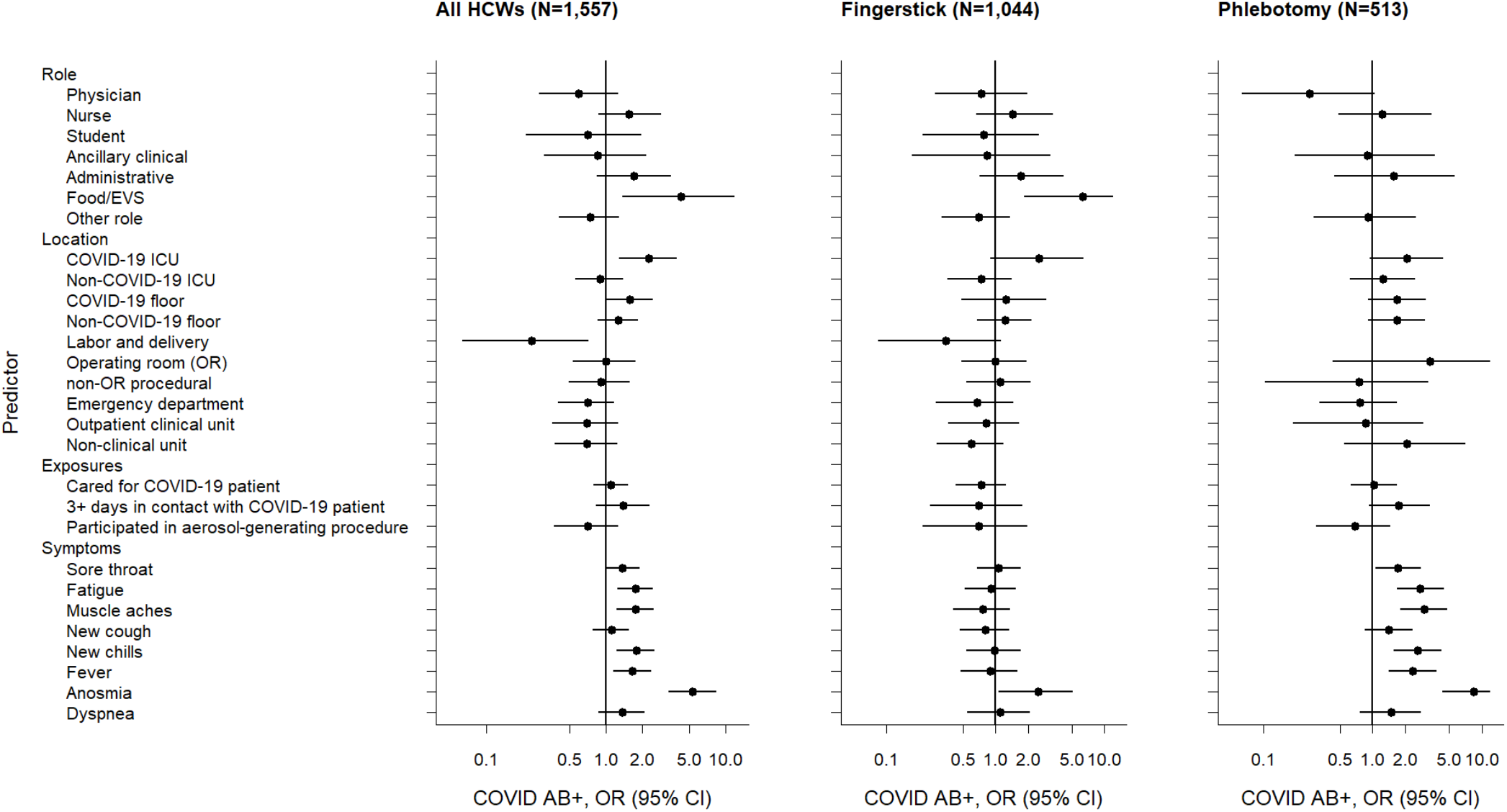
Forest plot of adjusted odds ratios (OR) of hypothesized predictors of COVID-19 seropositivity (AB+) among HCW study population and subgroups segregated by enrollment group and sample collection method (Fingerstick = open enrollment cohort; Phlebotomy = targeted enrollment cohort). ORs are adjusted for sex, age, race/ethnicity, known COVID-19 exposure outside of work, role, location, and COVID-19 patient contact. (EVS, environmental services)

## Appendices

### Appendix A. Survey utilized in this study

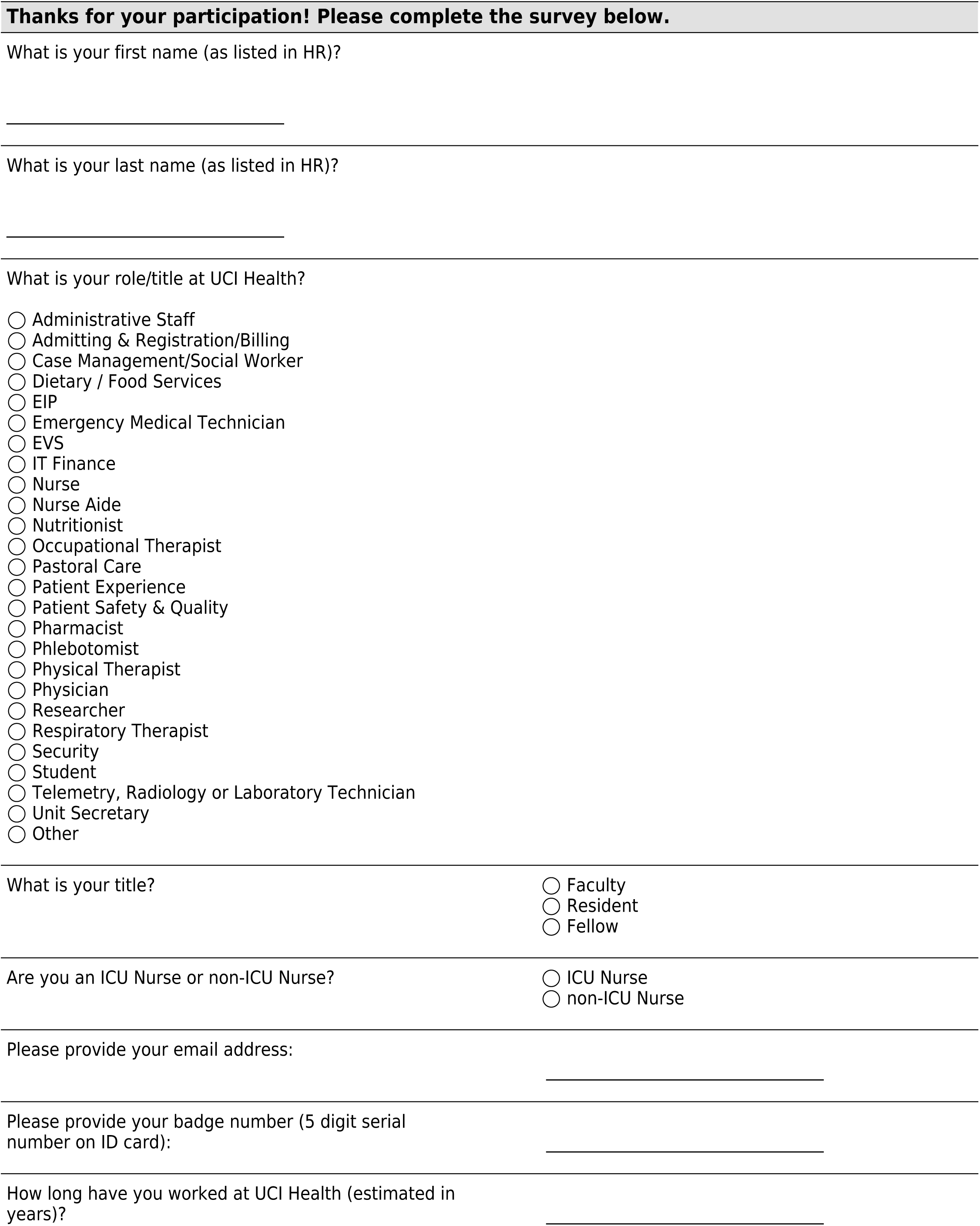
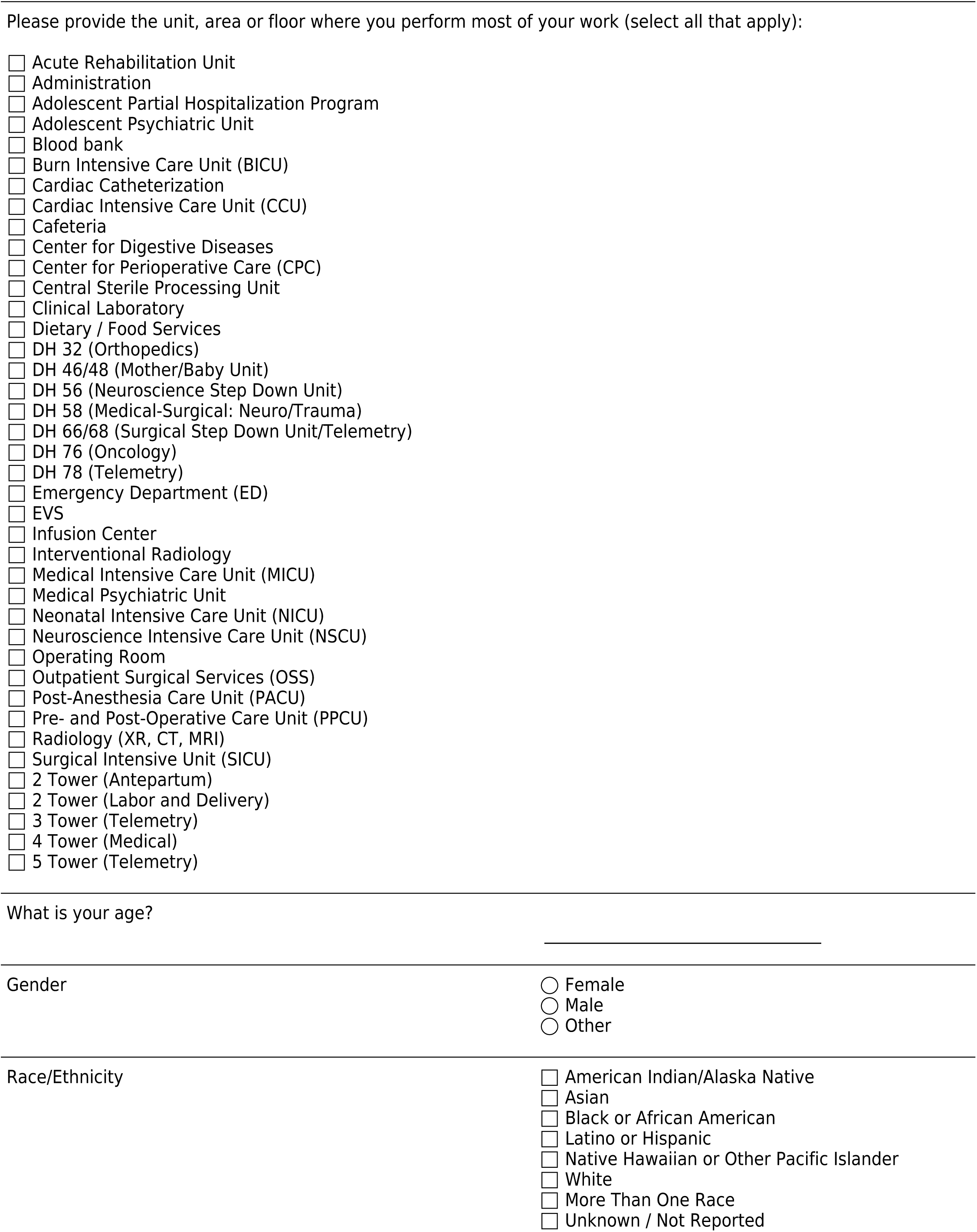
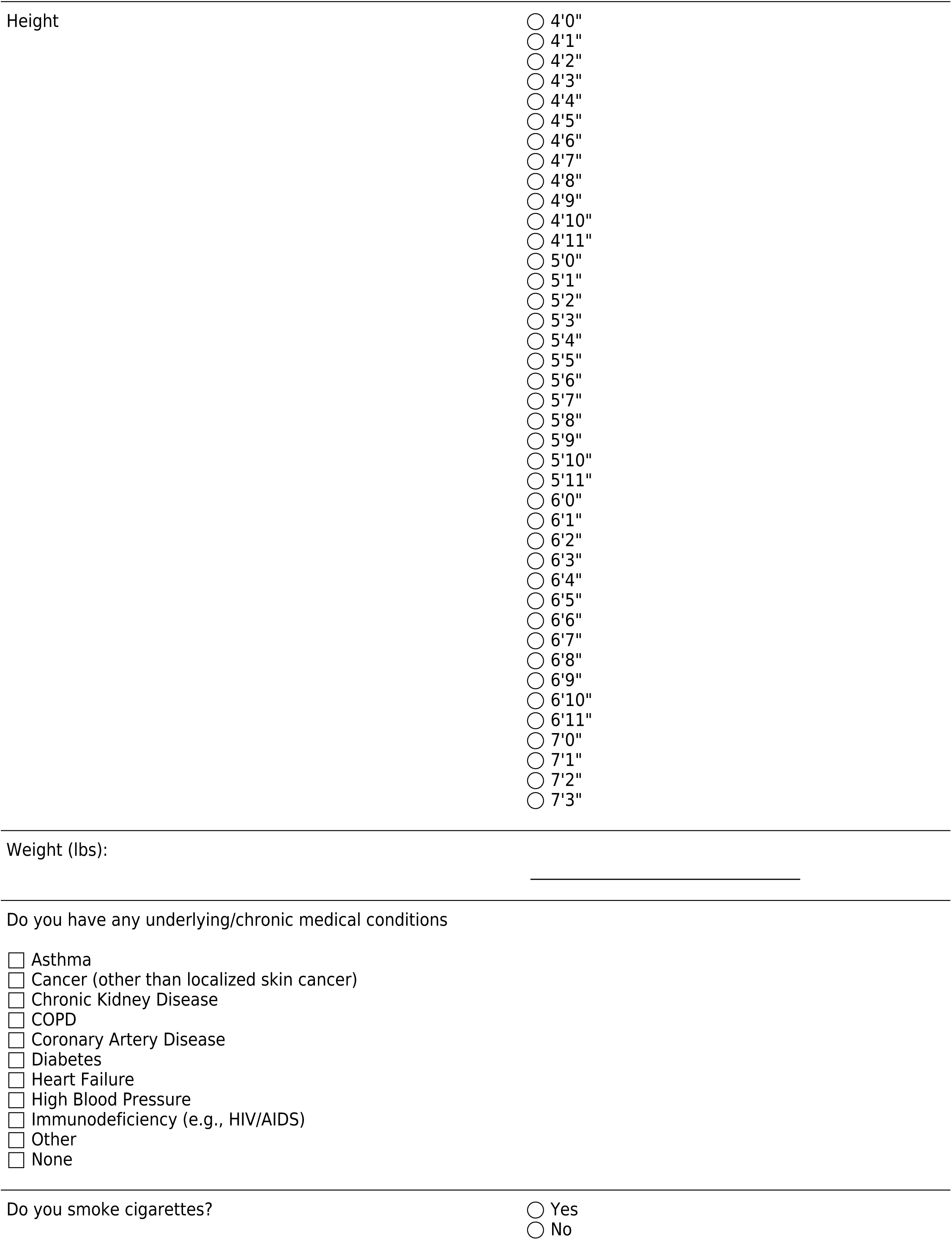
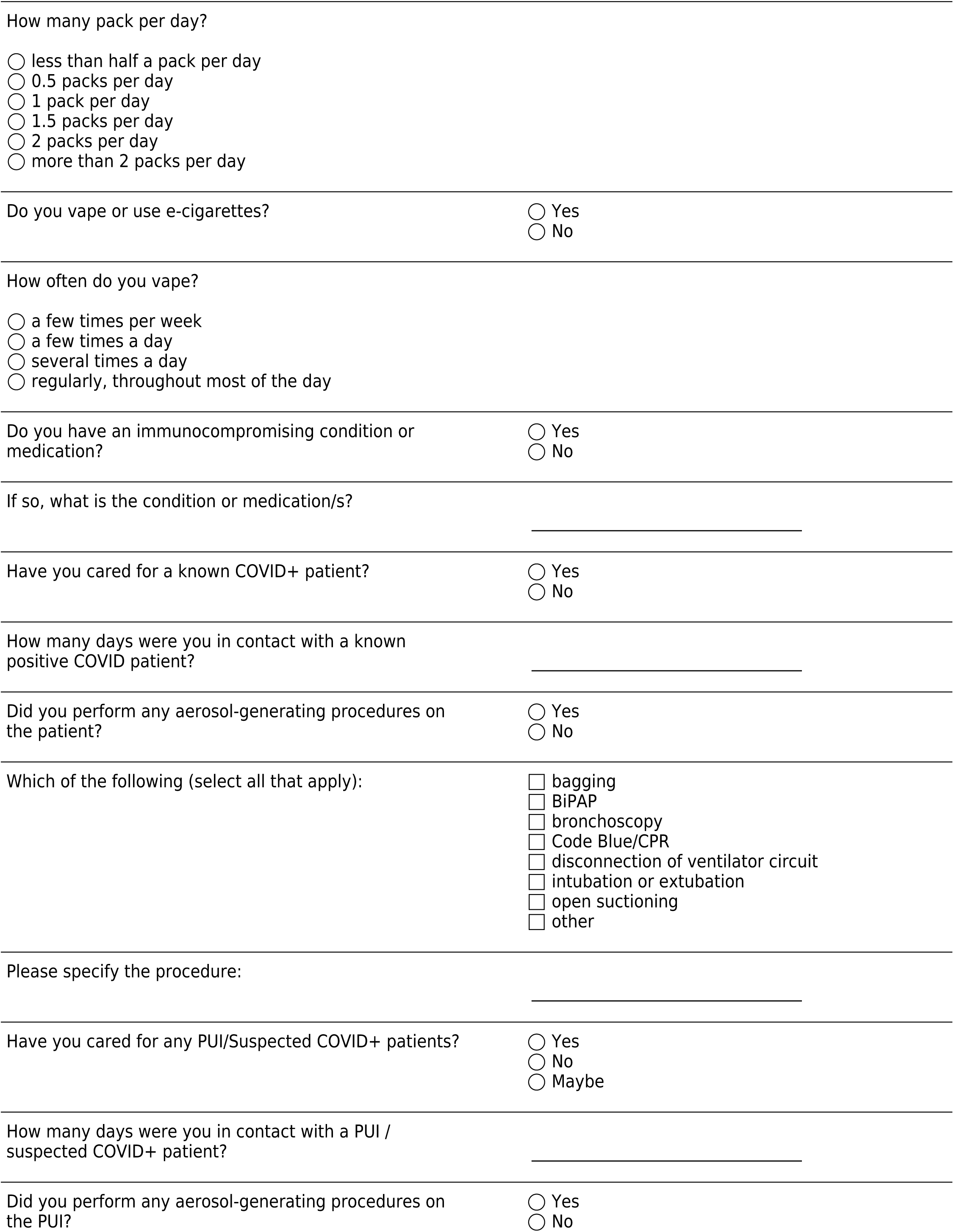
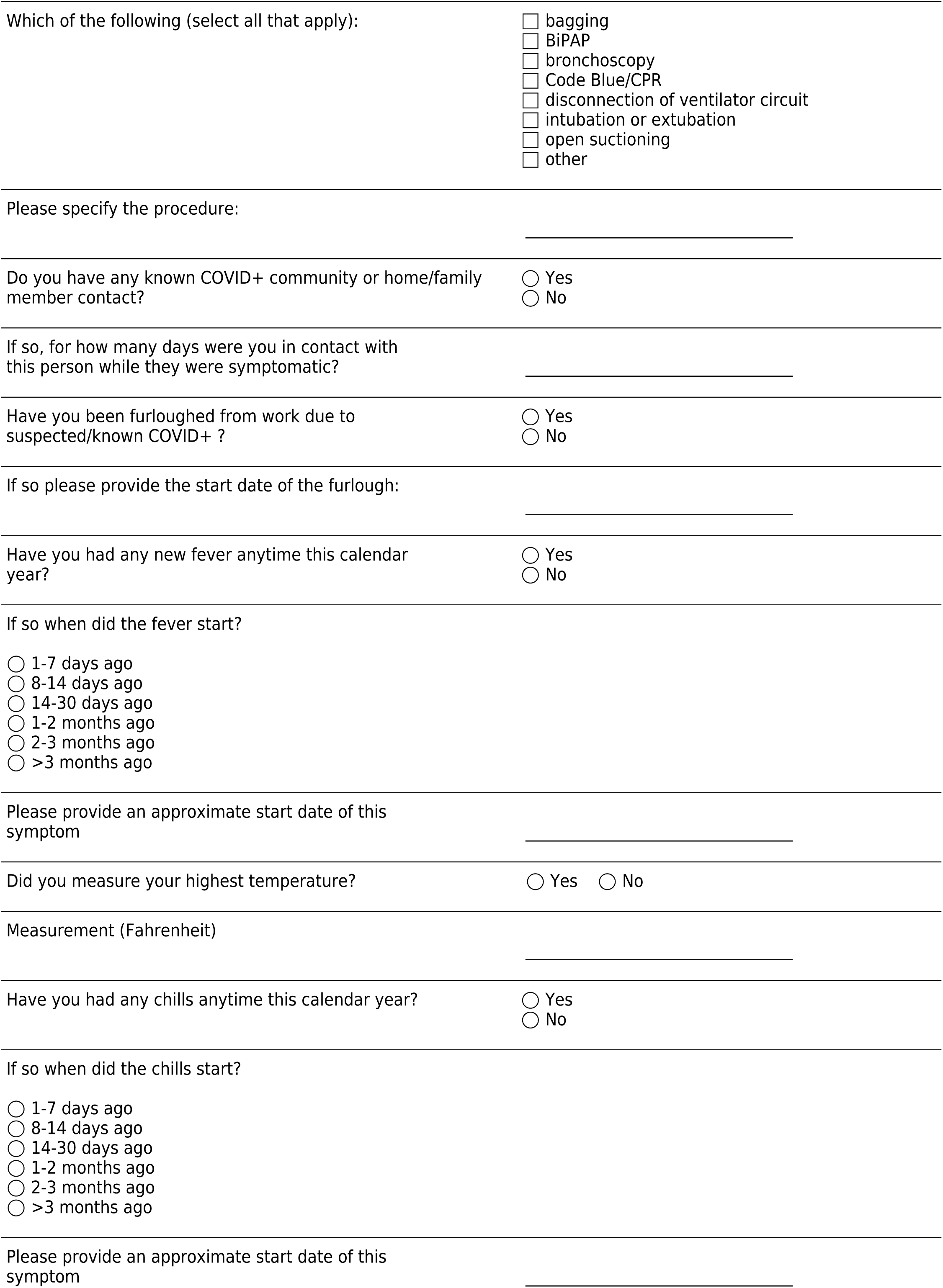
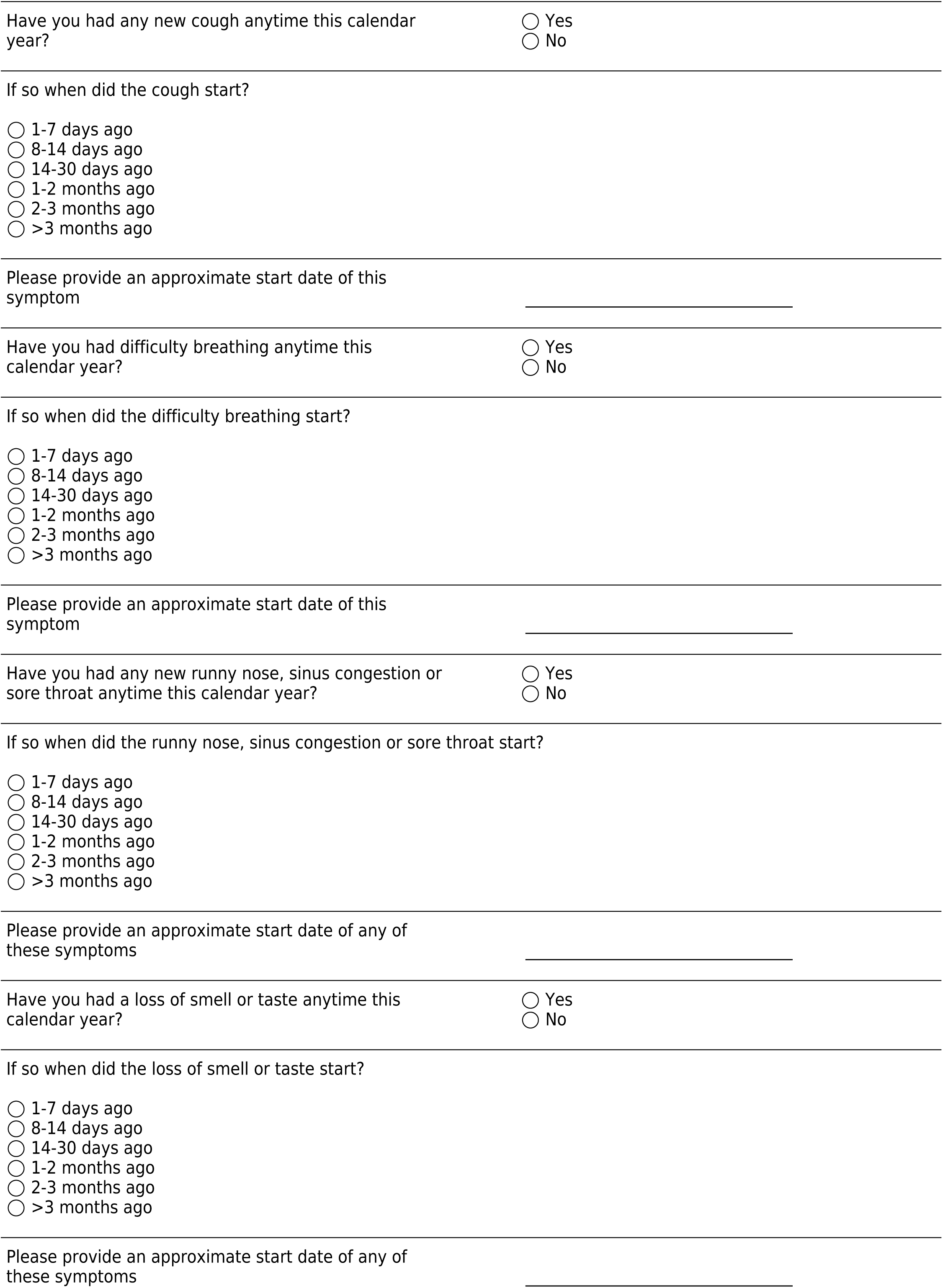
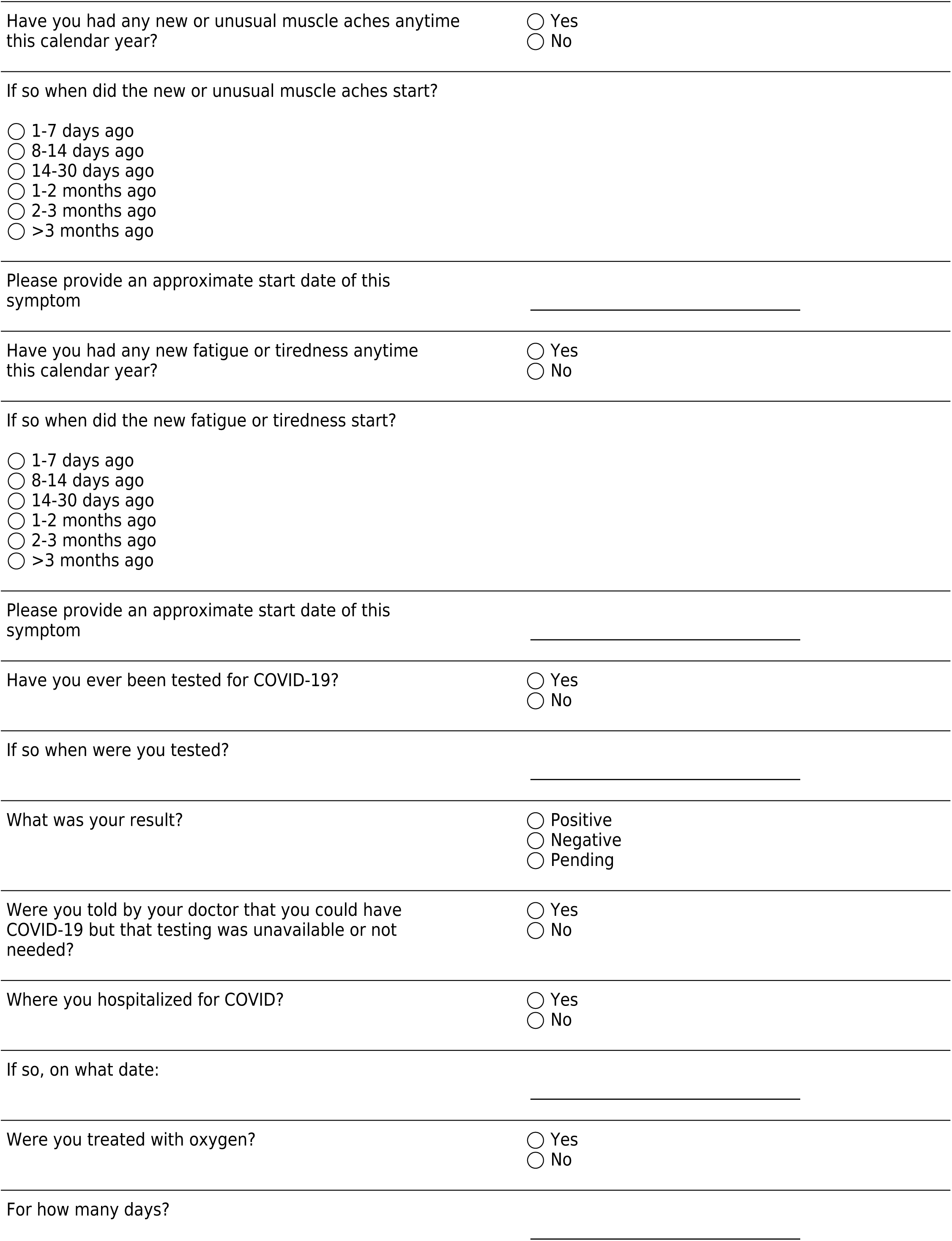
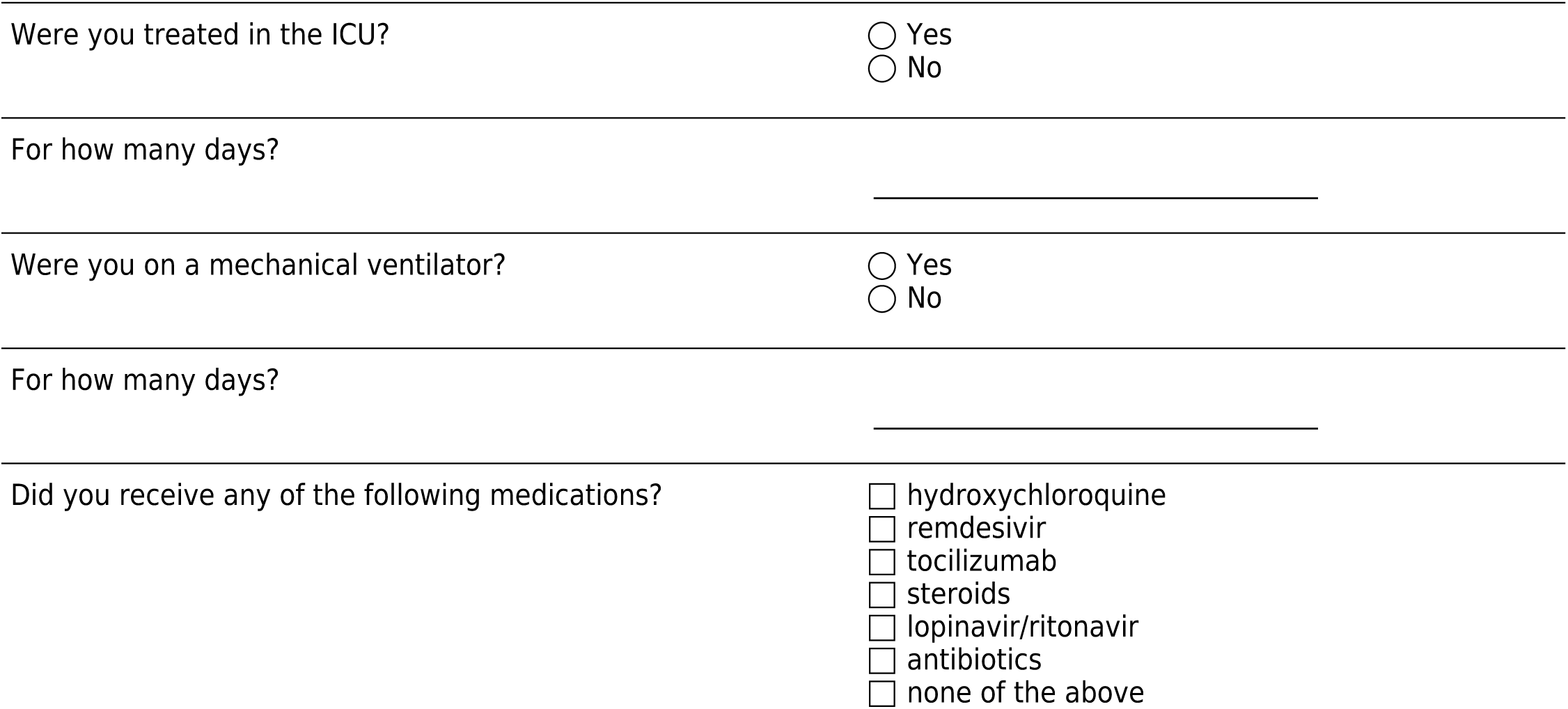

